# Differential immune response induced by two immunization schedules with an inactivated SARS-CoV-2 vaccine in a randomized phase 3 clinical trial

**DOI:** 10.1101/2022.08.05.22278464

**Authors:** Nicolás MS Gálvez, Gaspar A Pacheco, Bárbara M Schultz, Felipe Melo-González, Jorge A Soto, Luisa F Duarte, Liliana A González, Daniela Rivera-Pérez, Mariana Ríos, Roslye V Berríos, Yaneisi Vázquez, Daniela Moreno-Tapia, Omar P Vallejos, Catalina A Andrade, Guillermo Hoppe-Elsholz, Carolina Iturriaga, Marcela Urzua, María S Navarrete, Álvaro Rojas, Rodrigo Fasce, Jorge Fernández, Judith Mora, Eugenio Ramírez, Aracelly Gaete-Argel, Mónica Acevedo, Fernando Valiente-Echeverría, Ricardo Soto-Rifo, Daniela Weiskopf, Alba Grifoni, Alessandro Sette, Gang Zeng, Weining Meng, CoronaVac03CL Study Group, José V González-Aramundiz, David Goldblatt, Pablo A González, Katia Abarca, Susan M Bueno, Alexis M Kalergis

## Abstract

**Background:** The development of vaccines to control the COVID-19 pandemic progression is a worldwide priority. CoronaVac^®^ is an inactivated SARS-CoV-2 vaccine approved for emergency use with robust efficacy and immunogenicity data reported in trials in China, Brazil, Indonesia, Turkey, and Chile.

**Methods:** This study is a randomized, multicenter, and controlled phase 3 trial in healthy Chilean adults aged ≥18 years. Volunteers received two doses of CoronaVac^®^ separated by two (0-14 schedule) or four weeks (0-28 schedule). 2,302 volunteers were enrolled, 440 were part of the immunogenicity arm, and blood samples were obtained at different times. Samples from a single center are reported. Humoral immune responses were evaluated by measuring the neutralizing capacities of circulating antibodies. Cellular immune responses were assessed by ELISPOT and flow cytometry. Correlation matrixes were performed to evaluate correlations in the data measured.

**Results:** Both schedules exhibited robust neutralizing capacities with the response induced by the 0-28 schedule being better. No differences were found in the concentration of antibodies against the virus and different variants of concern between schedules. Stimulation of PBMCs with MPs induced the secretion of IFN-γ and the expression of activation induced markers for both schedules. Correlation matrixes showed strong correlations between neutralizing antibodies and IFN-γ secretion.

**Conclusions:** Immunization with CoronaVac^®^ in Chilean adults promotes robust cellular and humoral immune responses. The 0-28 schedule induced a stronger humoral immune response than the 0-14 schedule.

**Funding:** Ministry of Health, Government of Chile, Confederation of Production and Commerce & Millennium Institute on Immunology and Immunotherapy, Chile.

**Clinical trial number:** NCT04651790.

**summary:** Two immunization schedules were evaluated for the inactivated SARS-CoV-2 vaccine, Coronavac^®^, with two doses of the vaccine separated by two or four weeks. We compared humoral and cellular immune responses, showing they are mostly similar, with differences in neutralization capacities.

## Introduction

The current coronavirus disease 2019 (COVID-19) pandemic is caused by severe acute respiratory syndrome coronavirus 2 (SARS-CoV-2) [1, 2], a virus described for the first time at Wuhan, China, in late December 2019. SARS-CoV-2 is already responsible for almost 534 million cases of infection and over 6.31 million deaths during the last two years [3]. Worldwide efforts to develop effective vaccines against this virus have led to 10 vaccine prototypes approved for emergency use by the WHO, and over 8 billion vaccines administered to humans to date [4]. Most approved SARS-CoV-2 vaccines rely on a single viral component, namely the Spike (S) protein or its receptor-binding domain (RBD), which could negatively impact the neutralizing capacities of antibodies induced if circulating variants of concern (VOC), such as the Delta and Omicron, mutate those sequences [4]. Whole virus inactivated platforms have been widely used throughout history to prevent diseases against other viruses [5]. In the case of SARS-CoV-2, these vaccines contain a broader diversity of antigens than just the S protein. Therefore, they might be more suited to protect against emerging circulating variants [5, 6].

CoronaVac^®^ is an inactivated SARS-CoV-2 vaccine approved by the WHO for emergency administration to humans and developed by Sinovac Life Sciences Co., Ltd. [7, 8]. Phase 1/2 trials in China confirmed that this vaccine induces a robust immune response against SARS-CoV-2 [9, 10]. These data led to the evaluation of this vaccine in phase 3 clinical trials in other countries, including Brazil, Indonesia, Turkey, and Chile. An efficacy of 83.5% was reported for CoronaVac^®^ in Turkey and an effectiveness of 87.5% in Chile to prevent hospitalization due to COVID-19 for healthy adults [11, 12].

In this article, we compare the immune response elicited in healthy Chilean adults immunized with two doses of CoronaVac^®^ separated by either two (0-14 schedule) or four weeks (0-28 schedule). Our results suggest that, although the neutralizing capacities of antibodies elicited by a 0-28 immunization schedule with CoronaVac^®^ in Chilean adults are more robust than those induced by a 0-14 schedule, overall, both immune responses are within the same magnitude.

## Results

### A 0-28 day immunization schedule with CoronaVac^®^ promotes higher seropositivity rates and GMT values of neutralizing antibodies than a 0-14 schedule

To evaluate the humoral immune response elicited after vaccination with two doses of CoronaVac^®^, separated by two or four weeks (0-14 day and 0-28 day schedules, respectively) the neutralizing capacities of circulating antibodies were evaluated. This was performed independently through sVNTs and cVNTs for the ancestral strain, and pVNTs (Fig.1 and Fig.S2). For both immunization schedules, samples from 130 volunteers were tested for sVNT, 372 volunteers for cVNT with the ancestral strain, and 94 for pVNT (Table 1). These techniques show a robust increase in arbitrary WHO international units (IU), GMT values, and seropositivity rates two and four weeks after the second dose for both immunization schedules. Remarkably, as seen in IU for the sVNT and GMT values for cVNT and pVNT, the 0-28 schedule showed increased neutralizing capacities two and four weeks after the second dose (Fig.1A&B and Fig.S2A). No differences in seropositivity rates between both schedules were detected for any of the assays evaluated (Fig.1C&D and Fig.S2B). We also evaluated differences in the neutralizing capacities of circulating antibodies between the two age groups indicated for all four techniques (Fig.S3). Both age groups had significantly increased GMT values at all times compared to preimmune samples, irrespective of the immunization schedule and the technique evaluated (Table 2). These results suggest that CoronaVac^®^ induces robust production of circulating antibodies with neutralizing capacities after immunization with either a 0-14 or a 0-28 schedule. Remarkably, the 0-28 schedule promotes higher seroconversion rates and GMT or IU values of these neutralizing antibodies than the 0-14 schedule, as determined by sVNT and pVNT.

**Figure 1.**
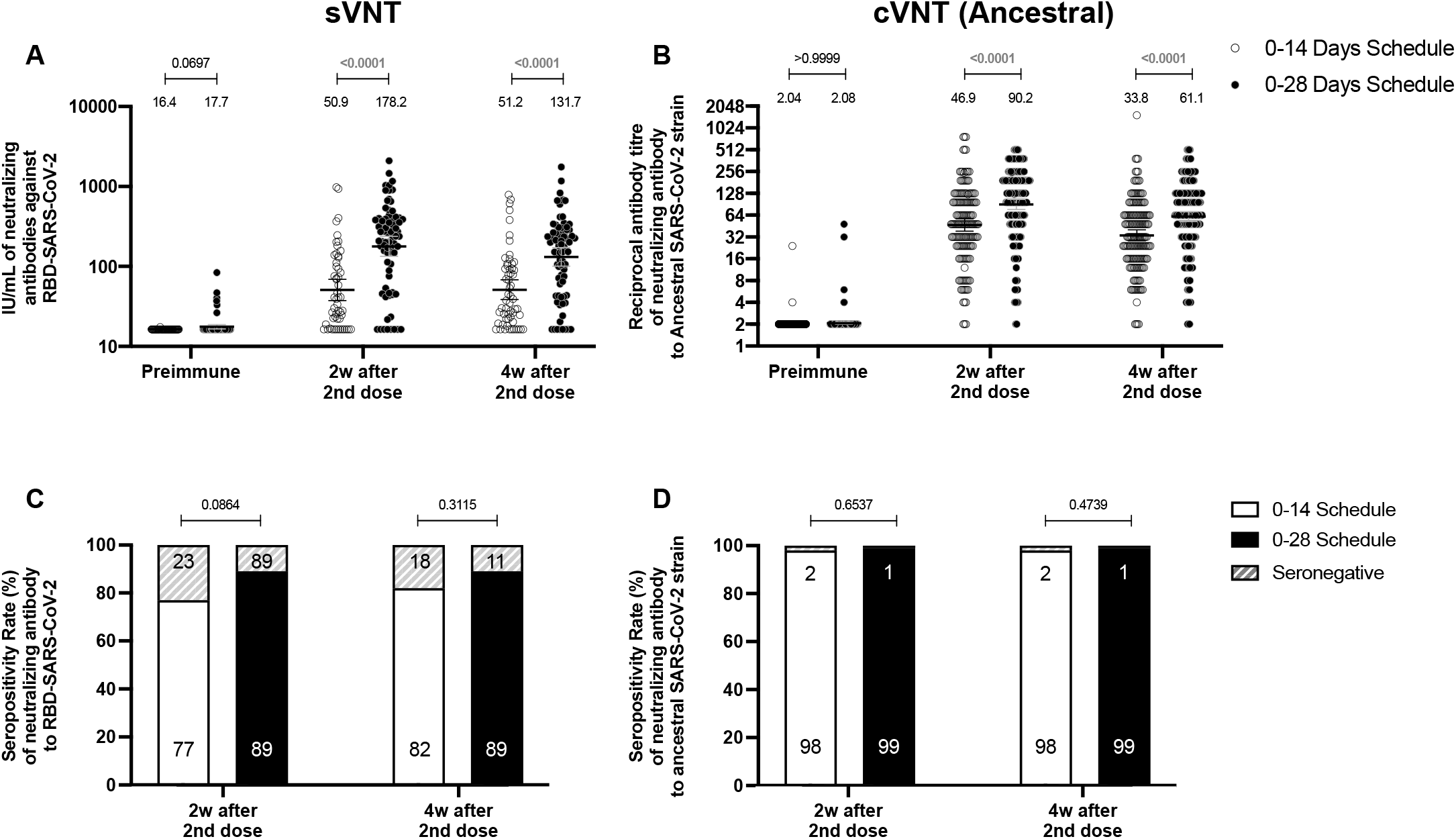
Circulating neutralizing antibodies against SARS-CoV-2 measured by sVNT and cVNT for the ancestral strain in immunized volunteers. Neutralizing antibody titers were evaluated with a surrogate virus neutralization test (sVNT), which quantifies the interaction between S1-RBD and hACE2 pre-coated on ELISA plates (**A,C**) and with a conventional plaque reduction neutralization test (cVNT), which quantifies the cytopathic effect (CPE) induced in Vero cells as plaques formation (**B, D**). Results were obtained from 372 volunteers for cVNT (ancestral) and 130 for sVNT for both schedules. Data is represented as the reciprocal antibody titer of neutralizing antibody v/s the different times evaluated. Numbers above the bars show either the arbitrary international units (IU) (**A**) or the Geometric Mean Titer (GMT) (**B**), and the error bars indicate the 95% CI. Seropositivity rates are also displayed (**C, D**). Data from IU and GMT values were analyzed by a two-tailed unpaired t-test of the base 2 logarithms of data to compare immunization schedules. Data from seropositivity rates were analyzed by a two-tailed Fisher’s exact test. Numbers above each bracket represent calculated P values comparing both immunization schedules.

**Table 1.**
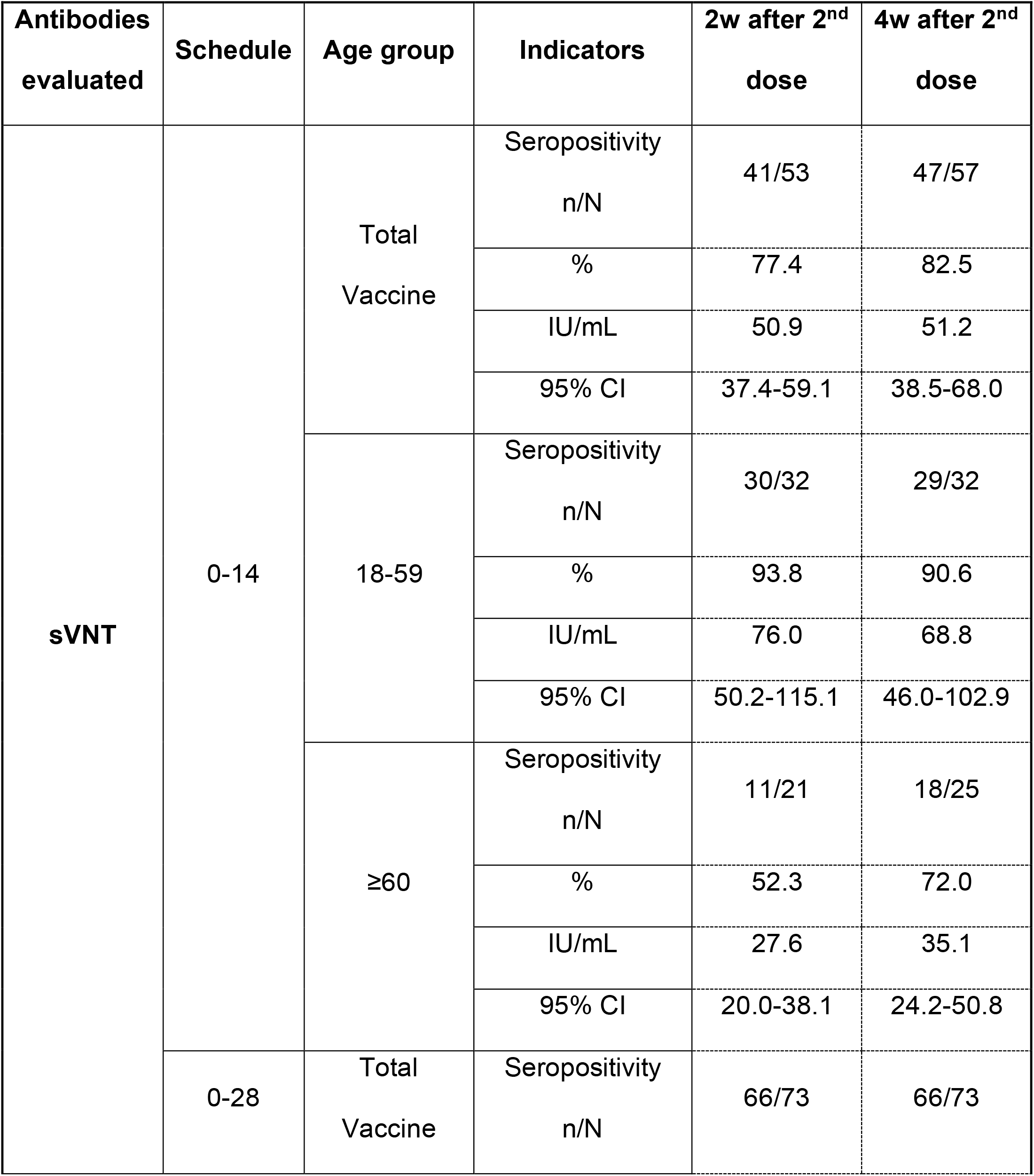
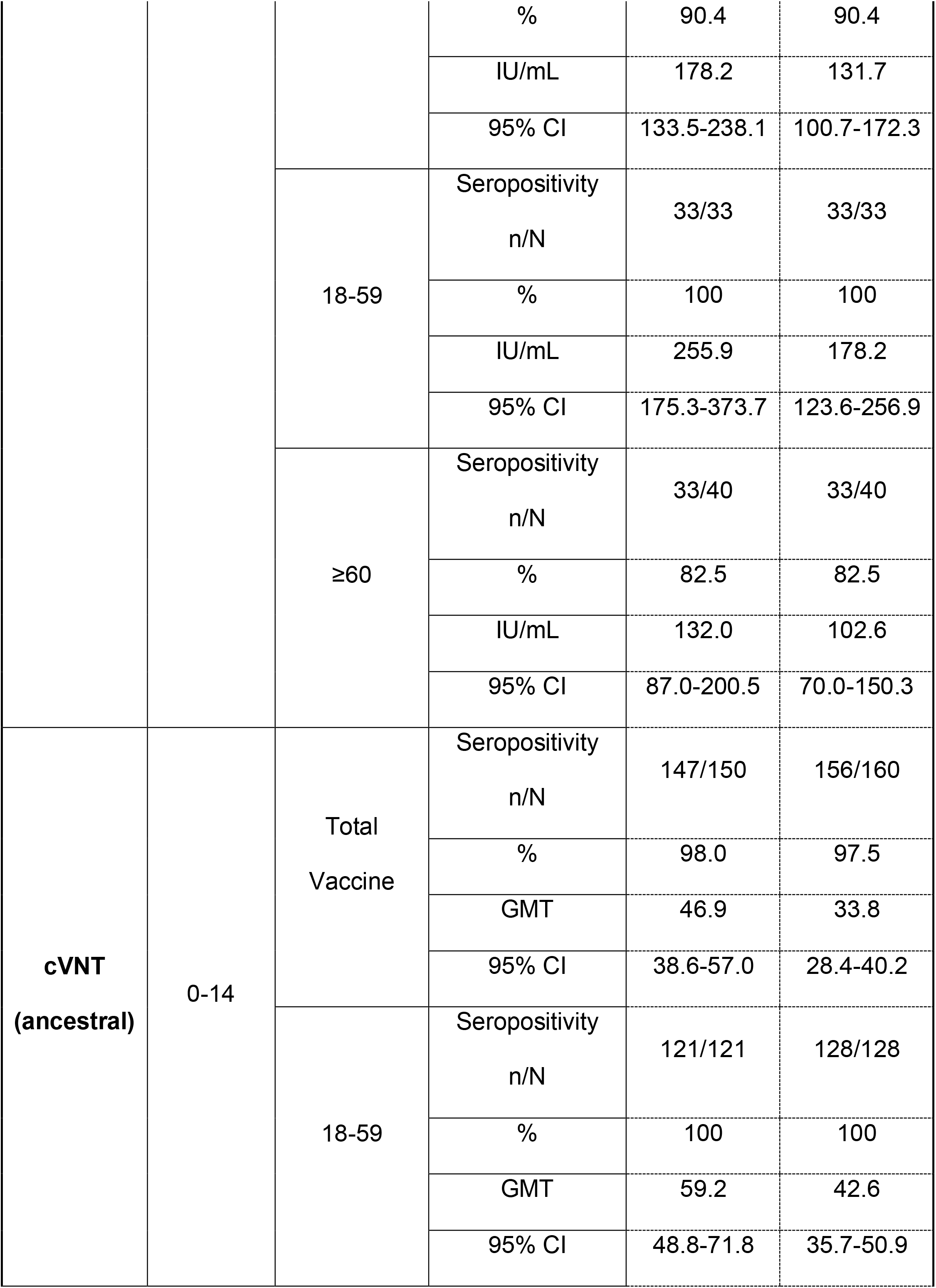
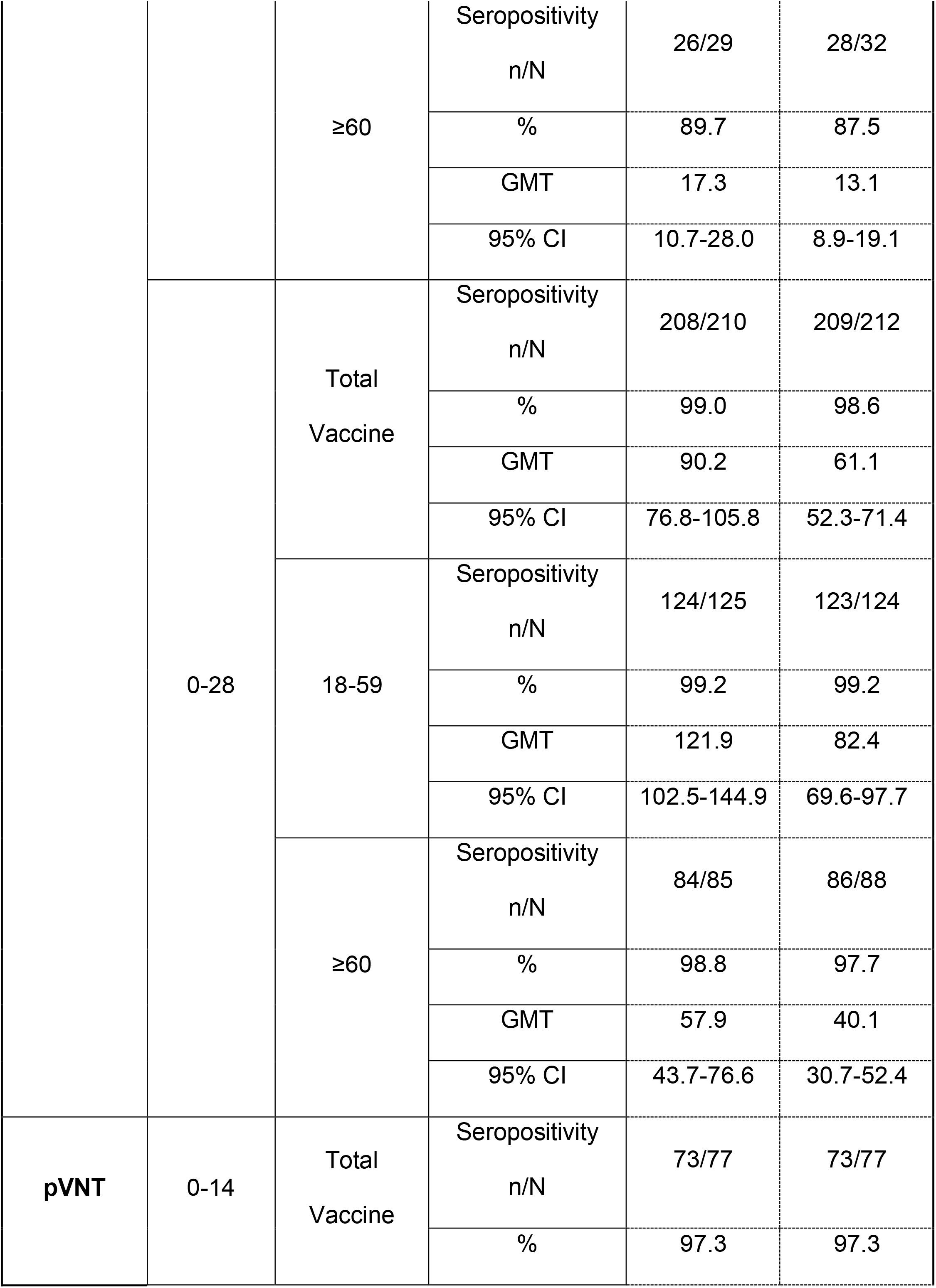
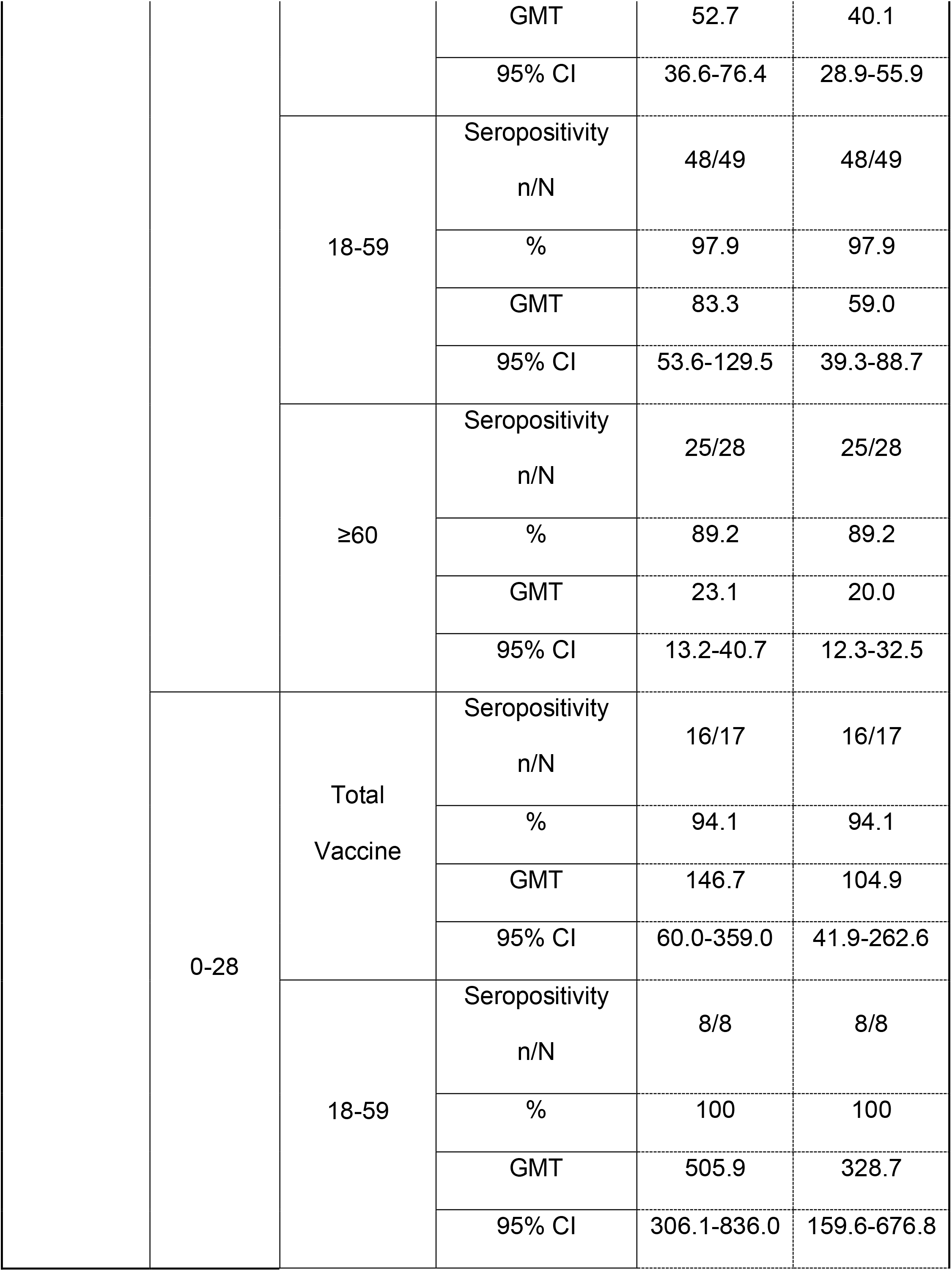
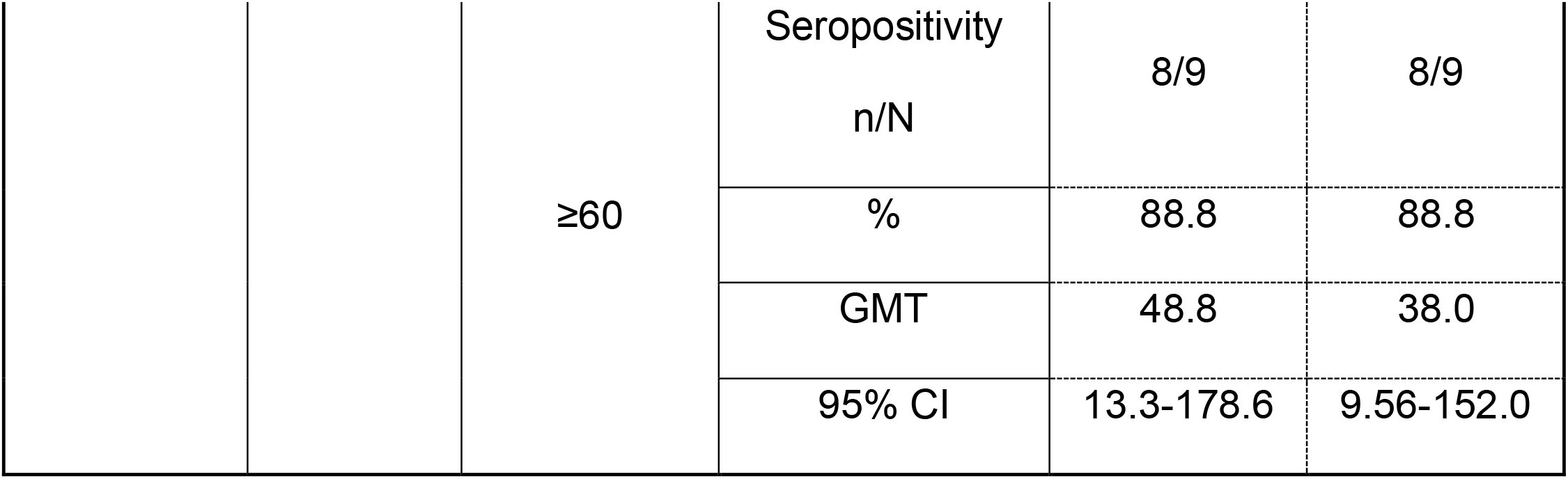
Seropositivity rates and GMT values measured for circulating neutralizing antibodies induced by CoronaVac^®^ in both immunization schedules and dissected by age group.

**Table 2.** P values estimated for neutralization assays evaluated for both immunization schedules.

| Figure | Parameter evaluated | Schedule;<br>Preimmune value | Preimmune compared to |  |
| --- | --- | --- | --- | --- |
|  |  |  | 2w after 2nd<br>dose | 4w after 2nd<br>dose |
| Figure<br>1A | sVNT<br>(IU/mL;<br>p value) | 0-14;<br>16.4 | 50.9;<br><0.0001 | 51.2;<br><0.0001 |
|  |  | 0-28;<br>17.7 | 178.2;<br><0.0001 | 131.7;<br><0.0001 |
| Figure<br>1B | cVNT (Ancestral)<br>(GMT;<br>p value) | 0-14;<br>2.04 | 46.9;<br><0.0001 | 33.8;<br><0.0001 |
|  |  | 0-28;<br>2.08 | 90.2;<br><0.0001 | 61.1;<br><0.0001 |
| Figure<br>S2 | pVNT<br>(GMT;<br>p value) | 0-14;<br>2.05 | 52.66;<br><0.0001 | 40.14;<br><0.0001 |
|  |  | 0-28;<br>2.0 | 146.71;<br><0.0001 | 104.93;<br><0.0001 |

\*P values were determined by performing One-way ANOVAs for repeated measures over Log<sub>10</sub> of data, followed by *post hoc* Bonferroni's multiple comparisons test. Red values indicate statistically significant results (p<0.05).

### Both immunization schedule with CoronaVac^®^ exhibits similar levels of anti-S1 and anti-RBD-specific antibodies

To evaluate the humoral immune response elicited after vaccination with two doses of CoronaVac^®^, separated by two or four weeks (0-14 day and 0-28 day schedules, respectively), antibody titers against the ancestral S1 and the RBD of SARS-CoV-2 were evaluated before administration of the first and second dose, and two and four weeks after the second dose (Fig.2). Samples from 162 volunteers were assessed independently for the S1-RBD through ELISA assays (Fig.2A) and 44 through meso-scale discovery (MSD) immunoassays (Fig.2B). Circulating antibodies against the S1-RBD were robustly increased for both immunization schedules at all times evaluated after administration of the first dose (preimmune), as determined by geometric mean units (GMU) values of the arbitrary WHO international standard. No differences were found at all times evaluated for anti-S1-RBD specific antibodies between both schedules (Fig.2A). Accordingly, no differences could be found between schedules for the MSD analyses performed, either for the S protein (left) or the RBD (right) of the ancestral strain of SARS-CoV-2 (Fig.2B). This suggests that CoronaVac^®^ induces a statistically significant increase in anti-S1 and anti-RBD antibodies after immunization with a 0-14 or a 0-28 schedule.

**Figure 2.**
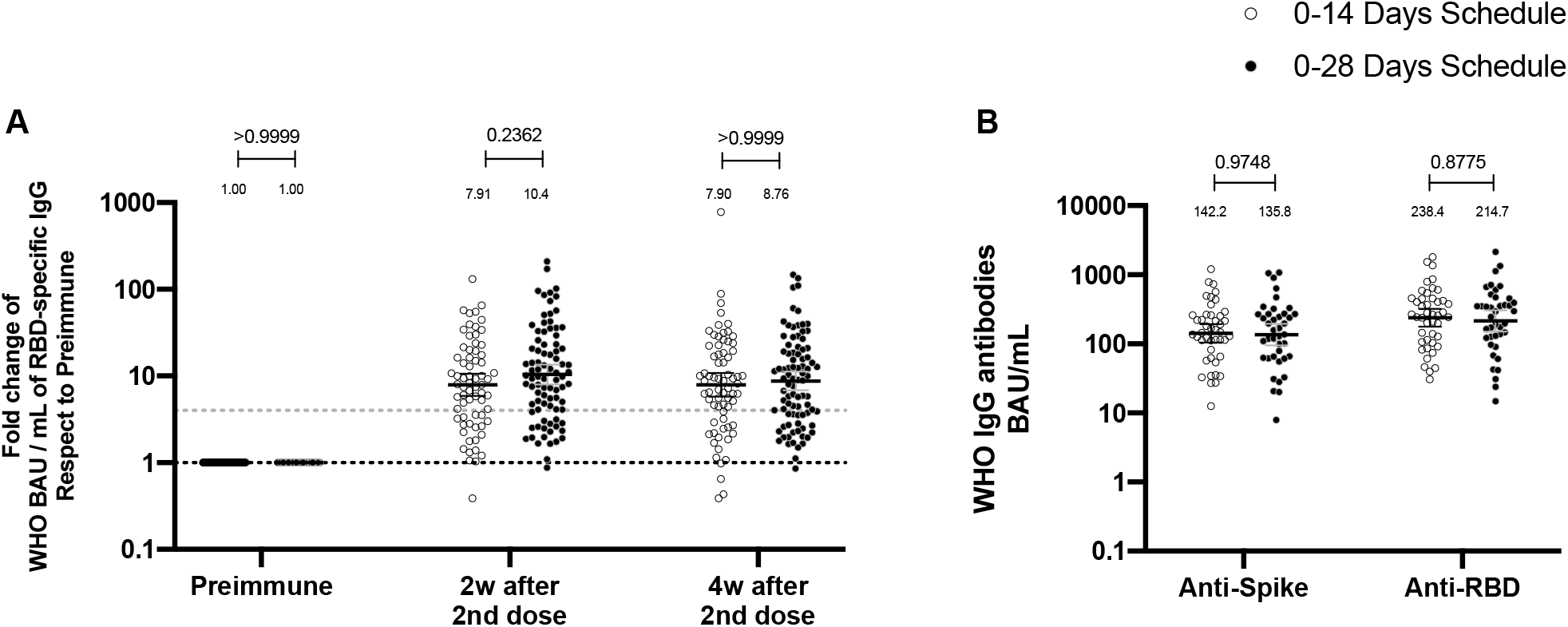
Total anti-S1 and anti-RBD antibodies circulating in immunized volunteers. Concentrations of IgG antibodies after two doses of CoronaVac^®^ were evaluated for immunized volunteers before the first (preimmune) and second dose and two and four weeks after the second. Specific IgG against the S1-RBD and the spike protein of SARS-CoV-2 were measured for 162 volunteers through ELISA assay (**A**) and 44 volunteers through MSD (**B**). Data are expressed as the reciprocal antibody titer in arbitrary WHO international unit v/s the different times evaluated. Error bars indicate the 95% CI. Spots represent individual values of each volunteer, with the numbers above each set of spots showing the GMU estimates. Data were analyzed using a two-tailed unpaired t-test of the Log_2_ of data to compare immunization schedules. Numbers above each bracket represent calculated P values comparing both immunization schedules. Dotted line on A is showing a value of 4, which is the threshold stablished for the seroconversion rate of each volunteer. Therefore, every spot over the dotted line represents volunteers that were considered positive for seroconversion relative to their preimmune sample.

### CoronaVac^®^ induces a significant cellular immune response against SARS-CoV-2 antigens regardless of the immunization schedule

To assess the cellular-mediated immune response elicited in volunteers immunized with CoronaVac^®^ in both immunization schedules, we evaluated the number of SFC positive for IFN-γ by ELISPOT and the expression of AIM on T cells by Flow Cytometry (Fig.3). PBMCs from 88 volunteers were evaluated for both immunization schedules and techniques. To evaluate SARS-CoV-2 antigen-specific secretion of IFN-γ and expression of AIM by T cells, PBMCs were stimulated independently with 4 MPs of peptides comprising the proteome of SARS-CoV-2. One MP contains peptides from the S protein (MP-S, 15-mer peptides), and another one considers the remaining viral proteome (MP-R, 15-mer peptides). The two other MPs comprise the whole proteome of SARS-CoV-2. These MPs were split in two as they were too many to be used as a single stimulus (MP-CD8A and MP-CD8B, 9-to 11-mer peptides). Stimulation of PBMCs with MP-S and MP-R induced a statistically significant compared to preimmune samples in the secretion of IFN-γ and the expression of AIM on CD4^+^ T cells (defined as OX40^+^ and CD137^+^) (Fig.S4A&S5A). This increase was not detected when stimulating with MP-CD8A and MP-CD8B (Fig.S4B&S5B). No statistical differences could be found between both immunization schedules in the total numbers of IFN-γ^+^ SFC (Fig.3A&B), or the percentage of AIM^+^ T cells (Fig.3C&D). No differences between both immunization schedules were found when evaluating the age groups indicated before for SFC (Fig.S6) or AIM on T cells (Fig.S7). No differences could be found between both immunization schedules for the number of IL-4^+^ SFCs (Fig.S8). Overall, these results indicate that immunization with CoronaVac^®^ in both schedules induces an increase in the number of IFN-γ^+^ SFC and the expression of AIM by CD4^+^ and CD8^+^ T cells upon stimulation with several MPs.

**Figure 3.**
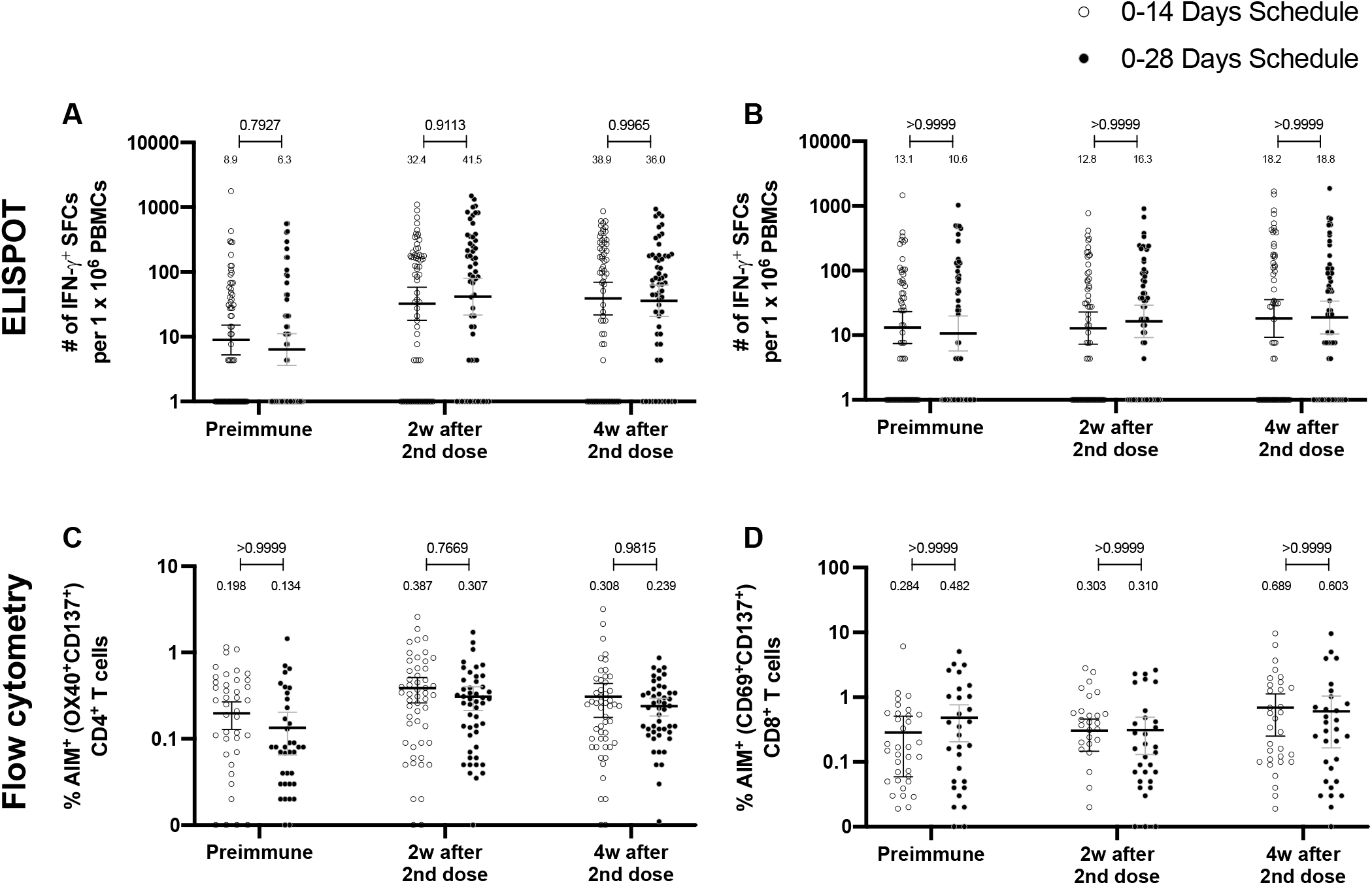
Similar levels of IFN-γ-secreting cells and expression of AIM on T cells are found upon stimulation with Mega Pools of peptides derived from SARS-CoV-2 for both immunization schedules with CoronaVac^®^. Total number of IFN-γ^+^ spot forming cells (SFC) were determined by ELISPOT. Data were obtained upon stimulation of PBMCs for 48 h with either MP-S or -R (**A**) and with either MP-CD8A or -B (**B**). The percentage of activated CD4^+^ (AIM^+^ [OX40^+^, CD137^+^]) and CD8^+^ (AIM^+^ [CD69^+^, CD137^+^]) T cells was determined by flow cytometry, upon stimulation for 24h with either MP-S or -R (**C**), and with either MP-CD8A or -B (**D**) in samples obtained before the first (pre-immune) and second dose, and two and four weeks after the second dose. A total of 124 samples were stimulated with MP-S and R for ELISPOT (**A**) and 116 for flow cytometry (**C**), and 117 with MP-CD8A and B for ELISPOT (**B**) and 110 for flow cytometry (**D**), for both schemes. Numbers above the bars show the mean and the error bars correspond to the 95% CI. Data were analyzed by a Mixed-effect two-way ANOVA, followed by a Bonferroni’s *post hoc* test to compare immunization schedules. Numbers above each bracket represent calculated P values comparing both immunization schedules.

### Immunization with CoronaVac^®^ induces a similar profile of antibodies against the S protein and the RBD of SARS-CoV-2 VOC regardless of the immunization schedule

To determine whether the immunization schedule had any impact on the profile of antibodies elicited against SARS-CoV-2 variants of concern (VOC), MSD immunoassays were performed to determine antibody titers against the S protein or the RBD from SARS-CoV-2 VOC (Fig.4). Samples from 44 volunteers in the 0-14 schedule and from 40 volunteers in the 0-28 schedule, obtained 4 weeks after the second dose, were evaluated. Anti-S antibodies against all variants tested (Alpha, Beta, Gamma, Delta, and Omicron) showed decreased concentrations compared to the Original strain regardless of the immunization schedule (Fig.S9A). A similar trend was observed for anti-RBD antibodies, although the concentration of antibodies against the Delta strain RBD seemed to remain similar to antibody levels against the Original strain RBD (Fig.S9B). The concentration of antibodies that recognize the Omicron RBD seemed to decrease in a more pronounced way compared to the decrease observed for antibodies that recognize the Omicron S protein. No differences were seen for the concentration of anti-S and anti-RBD antibodies between both schedules, when both age groups were evaluated together (Fig.4A&B). When both age groups were analyzed independently, no differences were seen in the 18-59 years group for anti-S and anti-RBD antibodies (Fig.S10A&B). For the >60 years group, decreased anti-S antibodies concentrations were found against the Alpha, Gamma, and Delta in the 0-28 schedule, compared to the 0-14 schedule (Fig.S11A). No differences in antibodies concentrations against the RBD were found for this age group, irrespective of the schedule (Fig.S11B).

**Figure 4.**
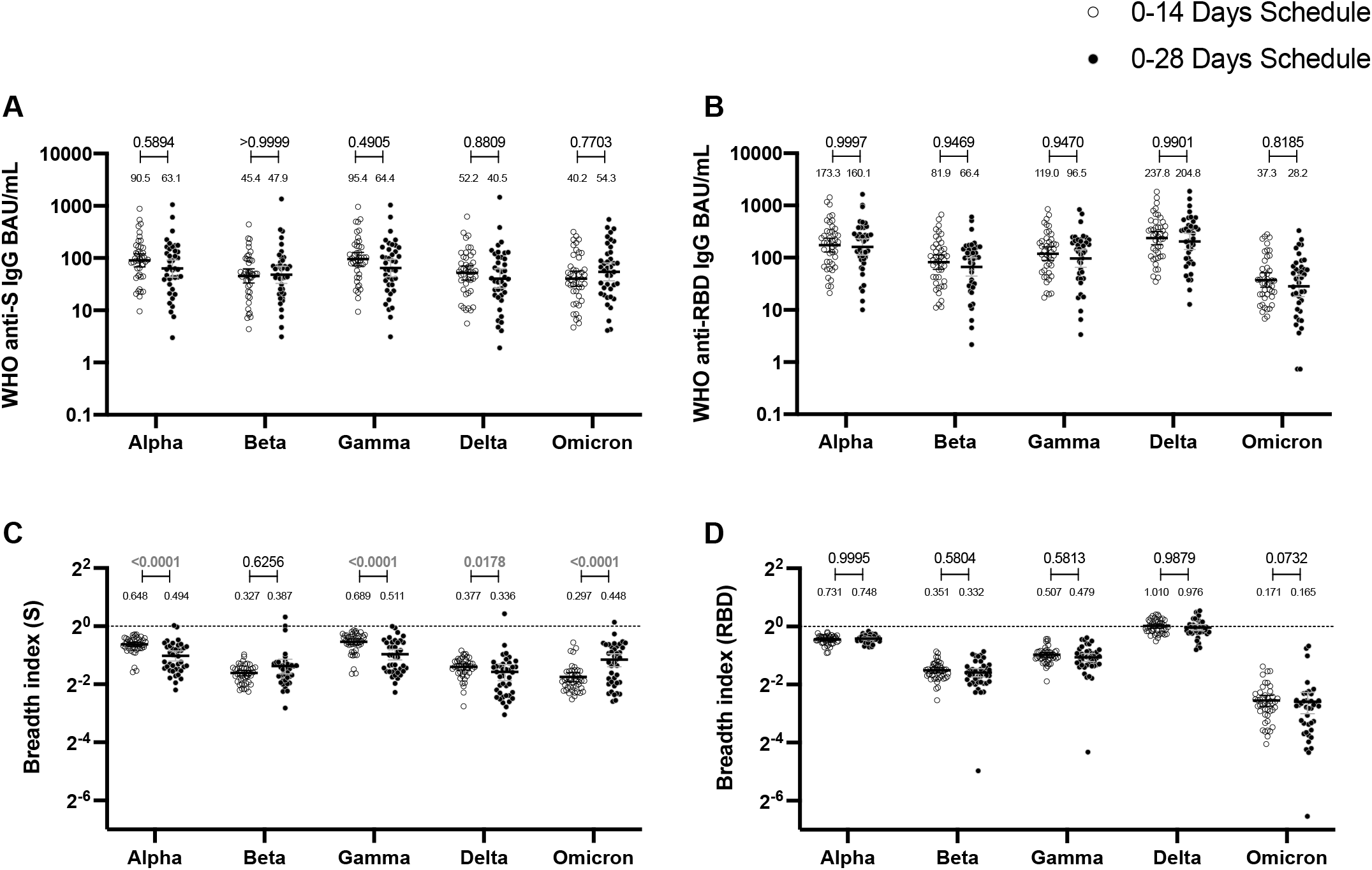
Antibodies against the S and RBD from VOC of SARS-CoV-2 are similar between schedules, while breadth index varies between schedules. Antibodies concentrations against the S (**A**) and the RBD (**B**) of different VOC of SARS were evaluated through MSD. Results were obtained from 44 volunteers for the 0-14 schedule and 40 volunteers for the 0-28 schedule, from samples obtained at 4 weeks after the second dose. Data is represented as the reciprocal antibody titer of neutralizing antibody v/s the different VOC evaluated. With these values, a breadth index was calculated for each VOC for anti-S (**C**) and anti-RBD (**D**) antibodies. Numbers above the bars show either the international units (IU) (**A, B**) or the breadh index (**C, D**), and the error bars indicate the 95% CI. Seropositivity rates are also displayed for all techniques (**D-F**). Data were analyzed by a Mixed-effect two-way ANOVA, followed by a Bonferroni’s *post hoc* test to compare immunization schedules. Numbers above each bracket represent calculated P values comparing both immunization schedules.

To account for differences on the antibody-binding activity of each volunteer’s serum, we calculated a Breadth Index, defined as the concentration of antibodies for a particular VOC divided by the concentration of antibodies for the Original strain. Differences in breadth of antibodies were found for the anti-S antibodies, with the 0-28 schedule showing decreased recognition capacity for Alpha, Gamma, and Delta, relative to the 0-14 schedule (Fig.4C). Interestingly, the 0-28 schedule showed increased recognition of the Omicron VOC, compared to the 0-14 schedule. We found no differences in this index for anti-RBD antibodies between schedules (Fig.4D). A decreased recognition capacity for all VOC of anti-S antibodies was detected (Fig.S10C). This was also seen in most VOC for anti-RBD antibodies, but no differences were found for the Delta strain (Fig.S10D). When analyzing the age groups, the 18-59 years group exhibited a reduced breadth index for the Alpha VOC in the 0-28 schedule, compared to the 0-14 schedule (Fig.S11C). No differences were seen for RBD (Fig.S11D). For the >60 years group, the 0-28 schedule exhibited a reduced breadth index for the Alpha, Gamma, and Delta VOC, compared to the 0-14 schedule. Remarkably, the 0-28 schedule reported increased breadth index for the Beta and Omicron VOC, relative to the 0-14 schedule.

These results suggest that CoronaVac can induce moderate cross-reactive humoral immune responses against SARS-CoV-2 VOC. While no evident patterns could be found between both schedules, the most marked differences can be detected when evaluating the >60 years old age group.

### The humoral and cellular immune responses elicited by both immunization schedules exhibit a significant correlation pattern

To identify potential correlations between variables, we generated correlation matrixes for each immunization schedule (Fig.5A&D). Overall, the neutralizing capacities determined by the different techniques exhibited a positive correlation value for both schedules. Particularly, the neutralizing capacities of circulating antibodies were strongly and positively correlated for the cVNT and sVNT evaluation at four weeks after the second dose (Fig.5B&E). These correlations were statistically significant and were found for both immunization schedules. Interestingly, positive correlations were also found for the neutralizing capacities of circulating antibodies as determined by cVNT two weeks after the second dose and the expression of AIM by CD8^+^ T cells four weeks after the second dose (Fig.5C&F). Again, these correlations were statistically significant and were found for both immunization schedules. These results suggest that the immune responses elicited by CoronaVac^®^ go hand in hand for either immunization schedule, as increased values of neutralizing antibodies are associated with increased expression of AIM by T cells.

**Figure 5.**
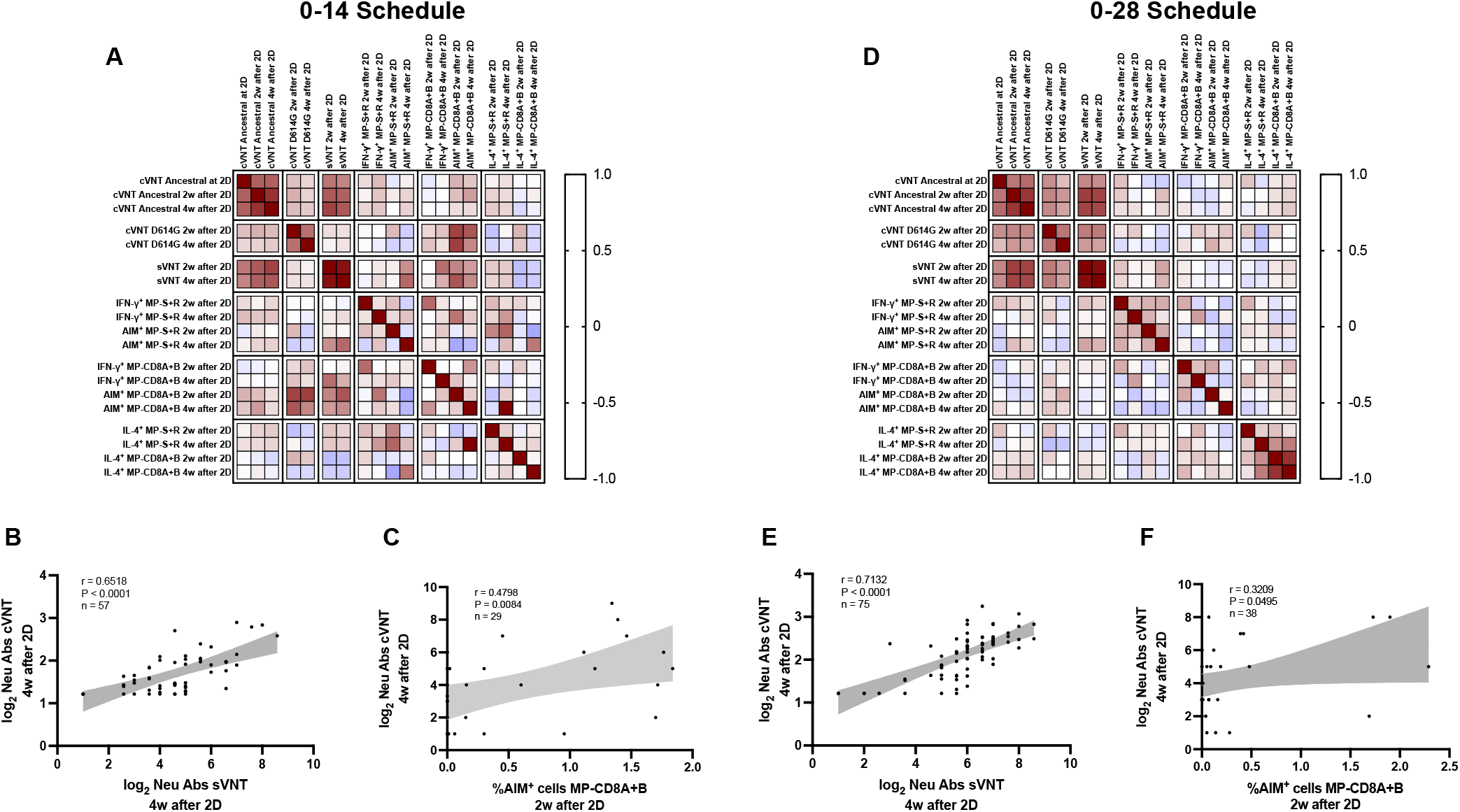
Multivariate analyses show correlated humoral and cellular immune responses. Pearson correlation matrixes were generated independently for the 0-14 (**A**) and 0-28 (**D**) immunization schedules, including humoral and cellular immune response variables. Colors indicate r values, and the scale is shown next to each matrix. Individual selected Pearson correlations for the 0-14 (**B-C**) and 0-28 (**E-F**) immunization schedules are shown, indicating n, r, and P values. Shaded gray areas show the 95% CI of the correlations.

## Discussion

Both SARS-CoV-2 and the ongoing COVID-19 pandemic have taken a considerable toll on the population worldwide and have posed a significant burden on the social well-being and the economies of developing and developed countries [3]. Scientific efforts have been directed towards generating safe and effective vaccines to prevent disease and severe cases associated with this virus, and the strive has not been vain [4]. Here, we report the immunogenicity profile elicited by healthy adults enrolled in a multicenter, randomized, and controlled phase 3 clinical trial performed with CoronaVac^®^, a whole virus inactivated SARS-CoV-2 vaccine manufactured by Sinovac Life Sciences, in Chile [7]. A total of 2,302 volunteers were enrolled in this Phase 3 trial, and a subset of these volunteers had their immunogenicity parameters evaluated. Circulating antibodies exhibited enhanced neutralizing capacities against SARS-CoV-2, as determined by four different assays. Differential responses antibodies concentrations against the S and the RBD of SARS-CoV-2 and different VOC were detected, with breadth indexes indicating varying degrees of changes against these strains. Increased secretion of IFN-γ and expression of activation-induced markers (AIM) were detected in T cells upon stimulation of peripheral blood mononuclear cells (PBMCs) with megapools (MPs) of peptides from SARS-CoV-2, as measured by ELISPOT and flow cytometry assays, respectively. We also performed multivariate correlation analyses to evaluate possible correlations between the measured parameters.

A prospective study considering a cohort of >10 million vaccinated persons in a 0-28 immunization schedule showed that the effectiveness of CoronaVac^®^ for this trial is 65.9% for preventing the development of COVID-19 symptoms, while a 90.3% was reported for the prevention of intensive care unit (ICU) admission [12]. Studies from Phase 1/2 clinical trials in China suggested that a 0-28 schedule may better induce a protective response against SARS-CoV-2 with CoronaVac^®^ [9,10,13]. Therefore, here we compared the immune response elicited by both schedules. Recent reports show that antibody titers against SARS-CoV-2 correlate with the protective capacities of COVID-19 vaccines [14]. Immunization with CoronaVac^®^ induced significantly increased levels of circulating neutralizing antibodies for both immunization schedules at all times after the first and second dose, irrespective of the age group evaluated. This is an expected response for vaccines that promote a protective response against SARS-CoV-2 and has also been reported for other vaccines such as BNT162b2 and mRNA-1273 [15].

The previous phase 1/2 trials held in China showed that a 0-28 immunization schedule is better at inducing anti-S1 and neutralizing antibodies in healthy adults, while in children and adolescents, it causes higher seroconversion rates before the second dose, compared to the 0-14 schedule [9, 16]. Here we show similar data regarding the neutralizing capacities of circulating antibodies, as arbitrary IU and GMT values were mainly increased two and four weeks after the second dose for the 0-28 schedule compared to the 0-14 schedule. It is important to note that the inactivated virus used in CoronaVac^®^ is the strain CZ02 [7], while the cVNT, sVNT, and pVNT use either the circulating ancestral and D614G strain, the S1-RBD, or the S protein from the original wild-type L strain [17], respectively. This could explain differences in neutralizing capacities reported among these techniques. Remarkably, no differences were found between schedules for the concentration of antibodies against the S protein and the RBD, at 4 weeks after the second dose. However, reduced breadth indexes were seen for the different VOC evaluated, with increased values against Beta and Omicron, but decreased values against the other variants in the 0-28 schedule, suggesting that a booster dose may be required to promote a more robust protective humoral immune responses against these emerging viruses [18].

Remarkably, it was recently shown that two doses of CoronaVac^®^ induce a steady cellular immune response against circulating variants of concern of SARS-CoV-2, while the neutralizing capacities of circulating antibodies were different among different strains [6]. This robust cellular response may be related to the presence of viral antigens other than the S protein of this virus and could be key when choosing vaccines to face this pandemic. The T cell immune responses elicited upon natural infection and vaccination are fundamental in modulating the disease caused by SARS-CoV-2 [19, 20]. Accordingly, several of the currently WHO-approved vaccine platforms have been shown to induce potent cellular immune responses, including those composed of recombinant proteins, mRNA, and viral vectors [21, 22]. The expression of AIM by T cells was mostly similar for both schedules at two and four weeks after the second dose, with no statistical differences. However, stimulation with MPs of 15-mer peptides induced an increased expression of AIM by CD4^+^ T cells relative to preimmune samples, two and four weeks after the second dose. The responses measured here indicate that a cellular response can be detected upon stimulation with these MPs initially selected *in silico* [23]. A Th1 response commonly associated with IFN-γ secretion is optimal for the clearance of intracellular pathogens, while Th2-related cytokines such as IL-4 may inhibit the polarization of CD4^+^ T cells towards this antiviral profile [9,10,13]. Although statistically higher than those detected for preimmune samples when stimulating with MPs of 15-mer peptides, numbers of IL-4^+^ SFC were remarkably lower than those seen for IFN-γ^+^ SFC. This is in line with the data previously reported for the 0-14 immunization schedule [13]. Our correlation matrixes showed that both immunization schedules could promote concerted humoral and cellular immune responses. Specifically, circulating neutralizing antibodies measured by different techniques were highly correlated two and four weeks after administration of the second dose. Positive and statistically significant correlations were found between neutralizing antibody titers determined by cVNT against the D614G variant two and four weeks after the second dose and the expression of AIM by CD8^+^ T cells four weeks after the second dose for both immunization schedules. This is especially important as both humoral and cellular immunity contribute to viral clearance, and vaccines should aim to develop both arms of the adaptive immunity [24].

This study also has caveats and limitations. Although the robust immunogenicity described here is encouraging, efficacy, hospitalization, and death prevention analysis are required to guide the use of this vaccine [9, 10]. Other limitations of this study are the lack of evaluation of long-term immunity (i.e., six or twelve months after the first dose), the partially homogeneous ethnicity of the evaluated population (healthy adults), a more exhaustive evaluation of cellular responses, and the immune response elicited against circulating variants of this virus.

Considering all the data presented in this article, we can conclude that immunization with CoronaVac^®^ in either a 0-14 or 0-28 vaccination schedule induces robust humoral and cellular responses in healthy adults from Chile. Further studies related to this phase 3 trial will be focused on the response elicited at later times after vaccination (i.e., six and twelve months after the first dose), the protection of CoronaVac^®^ towards circulating SARS-CoV-2 variants, and the capacity of a third dose to induce a robust immune response.

## Materials and methods

### Study design, randomization, masking, and volunteers

This clinical trial (ClinicalTrials.gov NCT04651790) was conducted in Chile at eight different sites, six located in Santiago city (Metropolitan Region) and two in the V Region of Valparaiso. The study protocol adhered to the current Tripartite Guidelines for Good Clinical Practices, the Declaration of Helsinki, and local regulations and was approved by the Institutional Scientific Ethical Committee of Health Sciences of the Pontificia Universidad Católica de Chile, (#200708006). The execution was approved by the Chilean Public Health Institute (#24204/20).

Recruited volunteers were adults aged ≥18 years, and informed consent was obtained upon enrollment. Volunteers received two doses of CoronaVac^®^ at day 0 and two (0-14) or four (0-28) weeks after the first immunization. Volunteers did not receive any payment for their participation. Study nurses oversaw the immunization and did not participate in any other study procedure. Inclusion and exclusion criteria were reported previously [13]. Exclusion criteria considered mainly history of confirmed symptomatic SARS-CoV-2 infection, pregnancy, allergy to vaccine components, and immunocompromised conditions. Well-controlled medical conditions were allowed.

Randomization was performed with a sealed enveloped system integrated with an electronic Case Report Forms (eCRF) in the OpenClinica platform. 2,302 volunteers were enrolled in this Phase 3 clinical trial by April 9th, 2021. 440 volunteers were part of the immunogenicity arm. 199 of these were part of the 0-14 schedule, and 241 were part of the 0-28 schedule (Fig.S1). The mean age of the recruited volunteers was 40.4±11.8 for the 0-14 schedule and 39.2±10.3 for the 0-28 schedule. A total of 52,9% volunteers were female and 47,1% were male.

CoronaVac consists of 3 µg or 600SU of β-propiolactone inactivated SARS-CoV-2 (strain CZ02) with aluminum hydroxide as an adjuvant in 0.5 mL [7]. Sodium chloride, monosodium hydrogen phosphate, and disodium hydrogen phosphate are excipients, and water for injection is included as solvent. A study nurse administered ready-to-use syringes with 0.5 mL of CoronaVac^®^ intramuscularly in the deltoid area. Sera and PBMCs were isolated from blood obtained before administration of the first and the second dose and two and four weeks after the second dose for both immunization schedules.

### Procedures

For the isolation of sera, 20 mL of blood were collected in anticoagulant tubes and distributed in 2 tubes of 10 mL per volunteer (BD Vacutainer Clot Activator tubes #367896). Blood was allowed to clot for at least 1h at RT. Samples were then centrifuged in a refrigerated centrifuge with a horizontal rotor at 1,300 x *g* for 10 min at 22°C. Serum was collected and stored at –80°C until use. Hemolyzed samples were rejected. For the isolation of PBMCs, blood was collected in 3 heparinized tubes (BD Vacutainer #367874, 10 mL) and stored at RT until processing. Samples were diluted with PBS (1:1) and centrifuged for 10 min at 1,200 x g (RT) in SepMate™ tubes (StemCell Technologies) with density-gradient medium (Lymphoprep). The plasma was then discarded and PBMCs were isolated by pouring them into a clean tube. Isolated PBMCs were washed twice with sterile PBS, counted, and cryopreserved in FBS (Industrial Biologicals) and 10% DMSO (Chem Cruz). All PBMC samples were stored in liquid nitrogen until use.

To assess the presence of anti-SARS-CoV-2 antibodies, blood samples obtained before the first dose (preimmune), before the second dose, two and four weeks after the second dose were analyzed. The quantitative measurement of human IgG antibodies against the RBD domain of the S1 protein (S1-RBD) was determined using the RayBio COVID-19 (SARS-CoV-2) Human Antibody Detection Kit (Indirect ELISA method) (Cat #IEQ-CoVS1RBD-IgG) and through meso-scale discovery (MSD) immunoassays were performed as described previously [25]. Briefly, these kits consist of 96-well plates coated with the S1-RBD protein segment SARS-CoV-2. Sera samples were serially diluted starting at a 200-fold dilution until a 6,400-fold dilution. After 1h of incubation at RT, the plates were washed, and a biotinylated anti-human IgG antibody provided in the kit was added and incubated for 30 min at RT. Plates were washed, and then an HRP-conjugated streptavidin was added and incubated for 30 min at RT. Plates were rewashed, and the TMB substrate solution supplied in the kit was added. Finally, a stop solution provided in the kit was added, and absorbance was measured at 450 nm in an ELISA plate reader (Biotek, Ref. 1506021). As controls, dilutions of the First WHO International Standard for anti-SARS-CoV-2 immunoglobulin (human - NIBSC code: 20/13), and positive control provided by each kit were included. Additional controls included samples of volunteers seropositive or seronegative at recruitment or inoculated with placebo. To calculate the end titers, the cut-off value for each test was determined as ≥2.1 times the average OD_450nm_ value of a panel of 8 serum samples from volunteers who received placebo and 12 seronegative serum samples. These serum samples were obtained before vaccination from seronegative volunteers. Seropositivity was determined as the highest dilution that reached >2.1 times the OD_450nm_ cut-off value. Seroconversion was defined as an increase of at least four times the titer at baseline.

For the surrogate virus neutralization test (sVNT), the SARS-CoV-2 Neutralizing Antibodies Test (BSNAT) Kit from BioHermes (Ref. COV-S41) was used to detect neutralizing antibodies in serum against SARS-CoV-2. This kit is a blocking assay which simulates the neutralization process, based on the ELISA platform. The main components of the kit are an ELISA plate pre-coated with the human ACE2 (hACE2) protein and the SARS-CoV-2 S1-RBD fragment conjugated with horseradish peroxidase (HRP-RBD). Briefly, the first step consisted in preparing serial dilutions from serum samples with the sample dilutor provided in the kit. Then, samples and controls were incubated with the HRP-RBD for 10 min at 37°C to allow the binding of neutralizing antibodies to RBD. Serums and controls previously incubated with HRP-RBD were added to ELISA plates (pre-coated with hACE2) and incubated for 20 min at 37°C. After incubation, samples were discarded, and all the wells were washed five times. Finally, TMB solution was added and quenched after 15 minutes of incubation at room temperature. Plates were read at 450 nm in a microplate reader (EPOCH, Biotek ref. 1506021). The neutralizing antibody titer was determined as the last fold-dilution with a cut-off value over 30% of inhibition. The inhibition rate was calculated based on the negative control absorbances (negative control – sample/negative control*100), and 10% was considered as the cut-off value.

For conventional virus neutralization test (cVNT), Vero E6 cells were infected with a SARS-CoV-2 strain obtained by viral isolation in tissue cultures (33782CL-SARS-CoV-2 strain). Neutralization assays were carried out by the reduction of cytopathic effect (CPE) in Vero E6 cells (ATCC CRL-1586). The titer of neutralizing antibodies was defined as the highest serum dilution that neutralized virus infection, at which the CPE was absent compared with the virus control wells (cells with CPE). Vero E6 cells (4×10^4^ cells/well) were seeded in 96-well plates. For neutralization assays, 100 µL of 33782CL-SARS-CoV-2 (at a dose of 100 TCID_50_) were incubated with serial dilutions of heat-inactivated sera samples (dilutions of 1:4, 1:8, 1:16, 1:32, 1:64, 1:128, 1:256, and 1:512) from volunteers for 1h at 37 °C. Then, the mix was added to the 96-well plates with the Vero E6 cells. Cytopathic effect on Vero E6 cells was analyzed seven days after infection. For each test, a serum sample from uninfected patients (negative control) and a neutralizing COVID-19 patient serum sample (positive control) were used.

For the pseudotyped virus neutralization test (pVNT), anti-SARS-CoV-2 neutralizing antibodies were measured using an HIV-1 backbone expressing firefly luciferase as a reporter gene and pseudotyped with the SARS-CoV-2 spike glycoprotein (HIV-1-SΔ19) as previously described [17]. Briefly, serum samples were initially diluted 1:4 in DMEM, serially diluted 1:3 up to 1:8,748, and then mixed with approximately 4.5 ng of p24 equivalents of HIV-1-SΔ19 in white 96-well plate. Plates were incubated for 1 hour at 37°C, and then 100 mL of DMEM containing 1×10^4^ HEK-ACE2 cells was added to each well. Firefly luciferase activity was measured 48h later, using the Luciferase Assay System (Promega) in a Glomax®-96 microplate luminometer (Promega). Estimation of the ID50 was obtained using a 4-parameter nonlinear regression curve fit measured as the percent of neutralization determined by the difference in average relative light units (RLU) between test samples and pseudotyped virus controls as previously described [17]. Data analyses and statistical analyses were carried out using GraphPad Prism v8.

To assess the cellular immune response, ELISPOT and flow cytometry assays were performed using PBMCs from volunteers at the different times indicated above. Upon thawing, cells were resuspended in fresh media in a 1:10 dilution to remove DMSO remnants from the freezing media. Then, cells were centrifuged, resuspended in fresh media, and counted in an automated cell counter (Logos Biosystems #L40001). Cells were adjusted to 6×10^6^ cells/mL and kept at 37°C, 5% CO_2_ for 15 min until use in the corresponding assay. ELISPOT plates containing a PVDF membrane were activated with 15 µL of 70% ethanol (Merck), washed three times with sterile 1x PBS, and then coated with human IFN-γ and IL-4 capture antibodies (1:250 and 1:125, respectively, CTL). After 3h of activation at RT, plates were washed two times with PBS and two times with PBS-Tween 20 0.05%. The stimulus included in these assays considers the use of Mega Pools (MPs) of peptides derived from SARS-CoV-2 proteins, previously described [23]. Two MPs composed of peptides from the S protein (MP-S) and the remaining proteins of the viral particle (MP-R) were used, as previously described [23]. These peptides were determined *in silico* to stimulate CD4^+^ T cells optimally. Also, two MPs composed of peptides from the proteome of SARS-CoV-2 (CD8-A and CD8-B) were used, as previously described [23]. These peptides were determined *in silico* to optimally stimulate CD8^+^ T cell. As positive controls, an independent stimulation performed with 5 mg/mL of Concanavalin A (ConA) (Sigma Life Science #C5275-5MG), and with an MP of peptides derived from cytomegalovirus proteins (MP-CMV) for the stimulation of both CD4^+^ and CD8^+^ T cells [23]. As a vehicle control, DMSO 1% (Merck #317275) was included. A total of 3×10^5^ cells in 50 µL of media were added to each well containing 50 µL of media with the corresponding stimulus. The final concentration of each stimulus per well was 1 µg/mL (except for ConA and DMSO). Positive controls for ELISPOT assays considered 5×10^4^ cells/well instead of 3×10^5^ cells/well. For ELISPOT assays, cells were incubated for 48h at 37°C, 5% CO_2_. After incubation, plates were washed one time with PBS, and three times with PBS-Tween20. Then, anti-human IFN-γ (FITC) and anti-human IL-4 (Biotin) antibodies (1:1,000 and 1:1,000, respectively) were added, and plates were incubated for 2h, RT. Plates were washed 3 more times with PBS-Tween 20 and then FITC-HRP and Streptavidin-AP (1:1,000) were added and plates were incubated for 1h, RT. After incubation, plates were washed 3 more times with PBS-Tween20. Then, plates were treated with the blue (15 min), and red (15 min) developer solution individually following the recommendations of the manufacturer. Plates were washed with tap water after each developer solution and allowed to dry for 24h prior to reading. To evaluate the number of T cells secreting IFN-γ, IL-4 or both, ELISPOT assays were performed with ImmunoSpot^®^ technology (ImmnunoSpot^®^ #hIFNgIL4-1M-10). Spot Forming Cells (SFCs) were counted on an ImmunoSpot^®^ S6 Micro Analyzer.

To characterize the expression of activation induced markers (AIM) by T cells, flow cytometry assays were performed. 3×10^5^ cells per well were stimulated as described for the ELISPOT assays, and after 24 h of incubation with the stimulus, samples were stained. Staining was performed by incubation for 45 min at 4°C using the following reagents: a mix of BD Horizon^TM^ Fixable Viability Stain 510 (BD Biosciences – CAT 564406 – 1 µL per 1×10^6^ cells); anti-CD14 V500 (BD Bioscience - clone M5E2); anti-CD19 V500 (BD Bioscience - clone HIB19); anti-CD3 AF-700 (Biolegend - clone OKT3); anti-CD69 PE (BD Bioscience - clone FN50); anti-CD8a BV-650 (BD Bioscience - clone RPA-T8); anti-CD4 BV-605 (BD Bioscience - clone RPA-T4); anti-CD137 (BioLegend, Clone 4-1BB); and anti-OX40 (BioLegend, Clone BER-ACT35). Cells were washed twice with 200 µL of PEB buffer, fixed, and then handed to the Flow Cytometry core facility, for their acquisition in an LSRFortessa X-20 flow cytometer.

### Statistical analyses

Sample size determination was already reported for this trial [13]. Statistical significance was set at α = 0.05 in all cases. Statistical analyses and symbols used for each analysis are described briefly in each figure legend. All statistical analyses were performed in GraphPad Prism v.9.0.1 or RStudio.

To evaluate statistical differences of anti-S antibody titers and neutralizing antibody titers either by cVNT, sVNT or pVNT induced by either immunization schedule, a two-tailed one-way ANOVA for repeated measures was performed over the Log_2_ of antibody titers, followed by Bonferroni’s multiple comparisons test in order to compare both schedules. Differences in seroconversion rates of neutralizing antibody titers were determined by a two-tailed Fisher’s exact test. Volunteers that were detected to be seropositive at entry for anti-N and/or anti-S antibodies were excluded from the analyses.

For the ELISPOT data, comparison between schedules was performed by a two-tailed one-way ANOVA for repeated measures was performed over the Log_2_ of antibody titers, followed by Bonferroni’s multiple comparisons test over the Log_10_ of the fold change. Differences among schedules for flow cytometry data were assessed by a two-tailed one-way ANOVA for repeated measures was performed over the Log_2_ of antibody titers, followed by Bonferroni’s multiple comparisons test of the percentage data.

Pearson correlation matrixes were generated for each immunization schedules considering both humoral and cellular immune response data. Humoral response data were transformed to the base 10 logarithm before analysis. Individual Pearson correlations were selected based on their immunological relevance, n, r, and P values. These values, as well as the 95% confidence bands of the correlation, are shown in each graph.

Other comparisons assessed were analyzed via two-tailed unpaired t tests, two-tailed non-parametric unpaired t tests (Mann-Whitney tests), two-tailed non-parametric paired t tests (Wilcoxon tests), ANOVA followed by Tukey tests, or two-tailed Fisher’s exact test, as indicated in the figure legends.

## Data Availability

All raw data (anonymized to protect the information of volunteers) is included with the publication of this article as a supporting file. Source Data File 1 contains the numerical data used to generate all the figures. The study protocol is also available online and was previously published in doi: 10.1101/2021.03.31.21254494.

## Acknowledgments

We would like to thank the Ministry of Health, Government of Chile; Ministry of Science, Technology, Knowledge, and Innovation, Government of Chile; and The Ministry of Foreign Affairs, Government of Chile and the Chilean Public Health Institute (ISP). We also would like to thank PATH for their support and sharing the First WHO International Standard for anti-SARS-CoV-2 immunoglobulin. We are grateful to Rami Scharf, Jessica White, Jorge Flores, and Miren Iturriza-Gomara from PATH for their support on experimental design and discussion; Alex Cabrera and Sergio Bustos from the Flow Cytometry Facility at Facultad de Ciencias Biológicas, Pontificia Universidad Católica de Chile for support with flow cytometry. We also thank the Vice Presidency of Research (VRI), the Direction of Technology Transfer and Development (DTD), the Legal Affairs Department (DAJ) of the Pontificia Universidad Católica de Chile. We are also grateful to the Administrative Directions of the School of Biological Sciences and the School of Medicine of the Pontificia Universidad Católica de Chile for their administrative support. Special thanks to the independent data safety monitoring committee (members in the Supplementary Appendix (SA)) for their oversight and to the subjects enrolled in the study for their participation and commitment to this trial. Members of the CoronaVac03CL Study Team are listed in the SA.

## Author contributions

**Conceptualization**: AMK, KA, SMB, PAG, JVG, GZ, WM, NMSG, GAP, BMS, FMG, JAS, LFD, LAG.

**Visualization**: AMK, KA, SMB, PAG, JVG, GZ, WM.

**Methodology**: NMSG, GAP, BMS, FMG, JAS, LFD, LAG, CI, MU, MSN, AR, RF, JF, JM, ER, AG-A, MA, FV-E, RS-R, GZ, WM, JVG-A, DG.

**Investigation**: NMSG, GAP, BMS, FMG, JAS, LFD, LAG, DR-P, MR, RVB, YV, DM-T, OPV, CAA, GH-E.

**Funding acquisition**: AMK, SMB.

**Project administration**: AMK, KA, SMB, PAG.

**Supervision**: AMK, SMB, PAG.

**Writing – original draft**: NMSG, GAP, BMS.

**Writing – review & editing**: AMK, SMB, PAG, NMSG, GAP, BMS, FMG, JAS, LFD.

**Verification of underlying data**: AMK, SMB, NMSG, GAP, BMS, FMG, JAS, LFD, LAG.

## Supplementary Figures Legends

**Figure S1.**
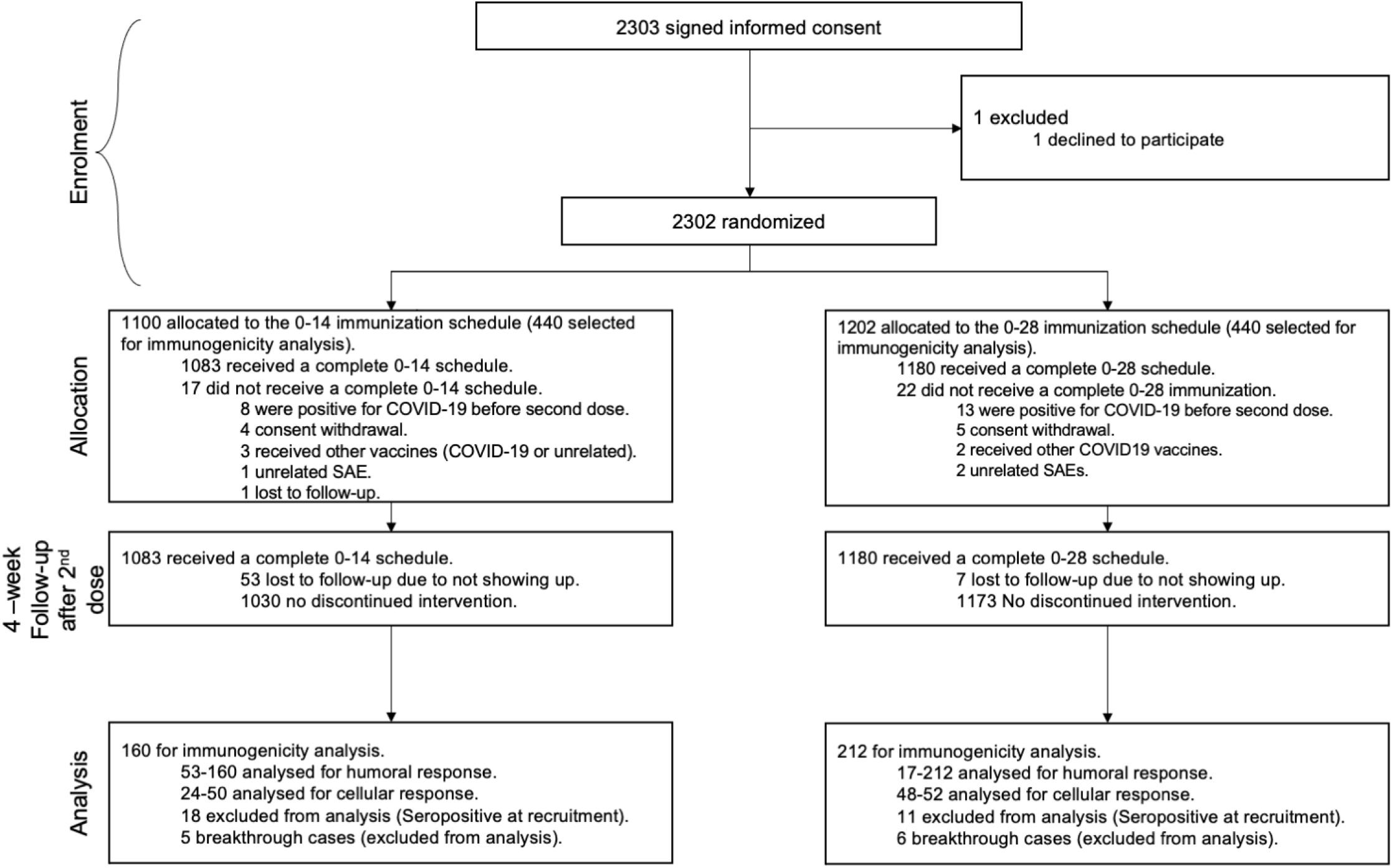
Study design for this Phase 3 clinical trial comparing two different immunization schedules as of August 2021. This study aims to characterize the differential immune response elicited by two immunization schedules with CoronaVac^®^, with each dose separated by either 2 or 4 weeks.

**Figure S2.**
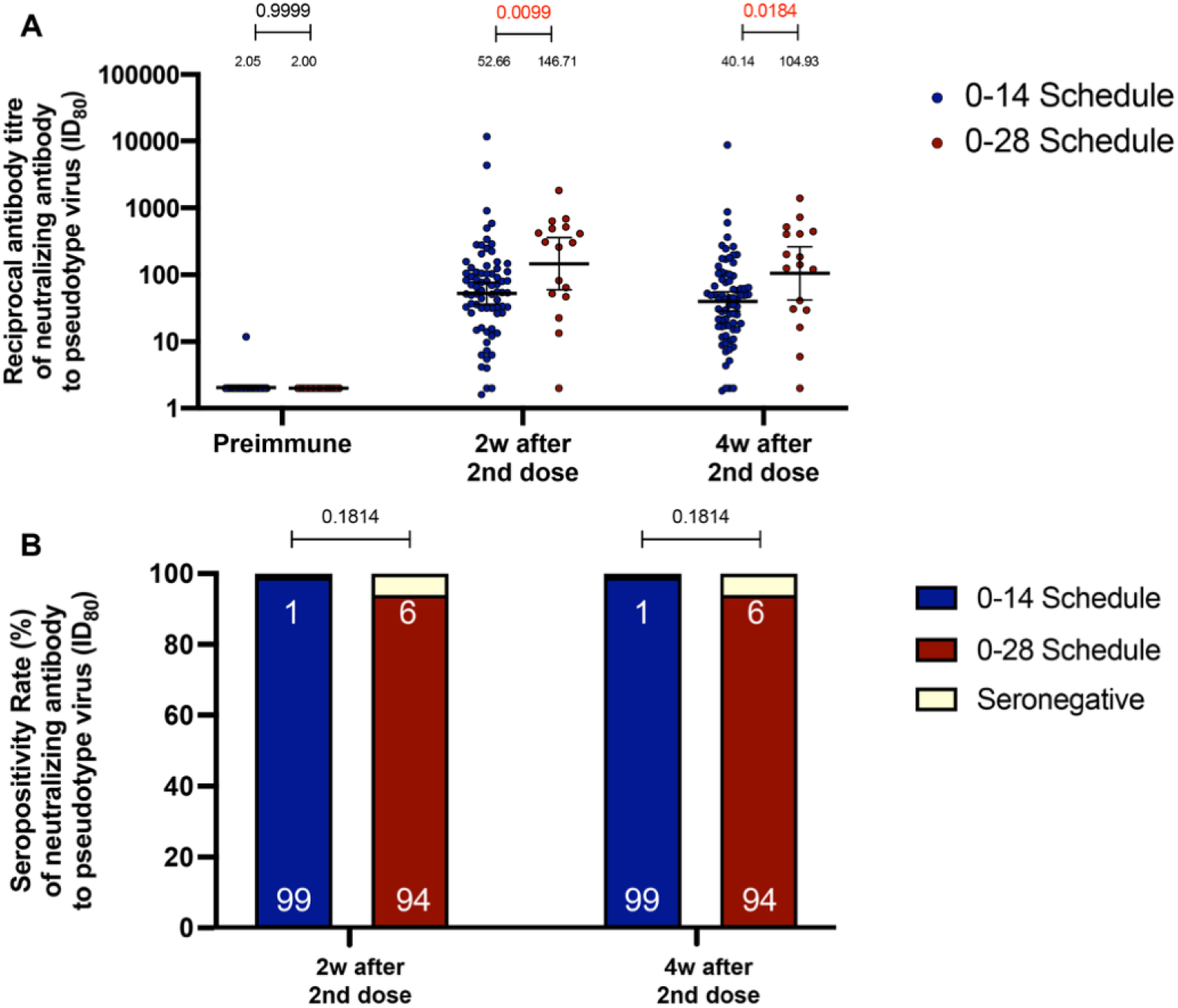
GMT values and seropositivity rates of circulating neutralizing antibodies against SARS-CoV-2 measured through pVNT (ID80). Neutralizing antibody titers were evaluated with a pseudovirus-based neutralization test (pVNT). Results were obtained from 94 volunteers for both schedules. In (**A**), data is represented as the reciprocal antibody titer of neutralizing antibody v/s the different times evaluated. Numbers above the bars show the Geometric Mean Titer (GMT), and the error bars indicate the 95% CI. A two-tailed, one-way ANOVA for repeated measures was performed over the Log_2_ of antibody titers, followed by Bonferroni’s multiple comparisons test to compare GMUs. (**B**) Data from seroconversion rates were analyzed by a two-tailed Fisher’s exact test. Values above the lines indicate P values. Significant P values are shown in red.

**Figure S3.**
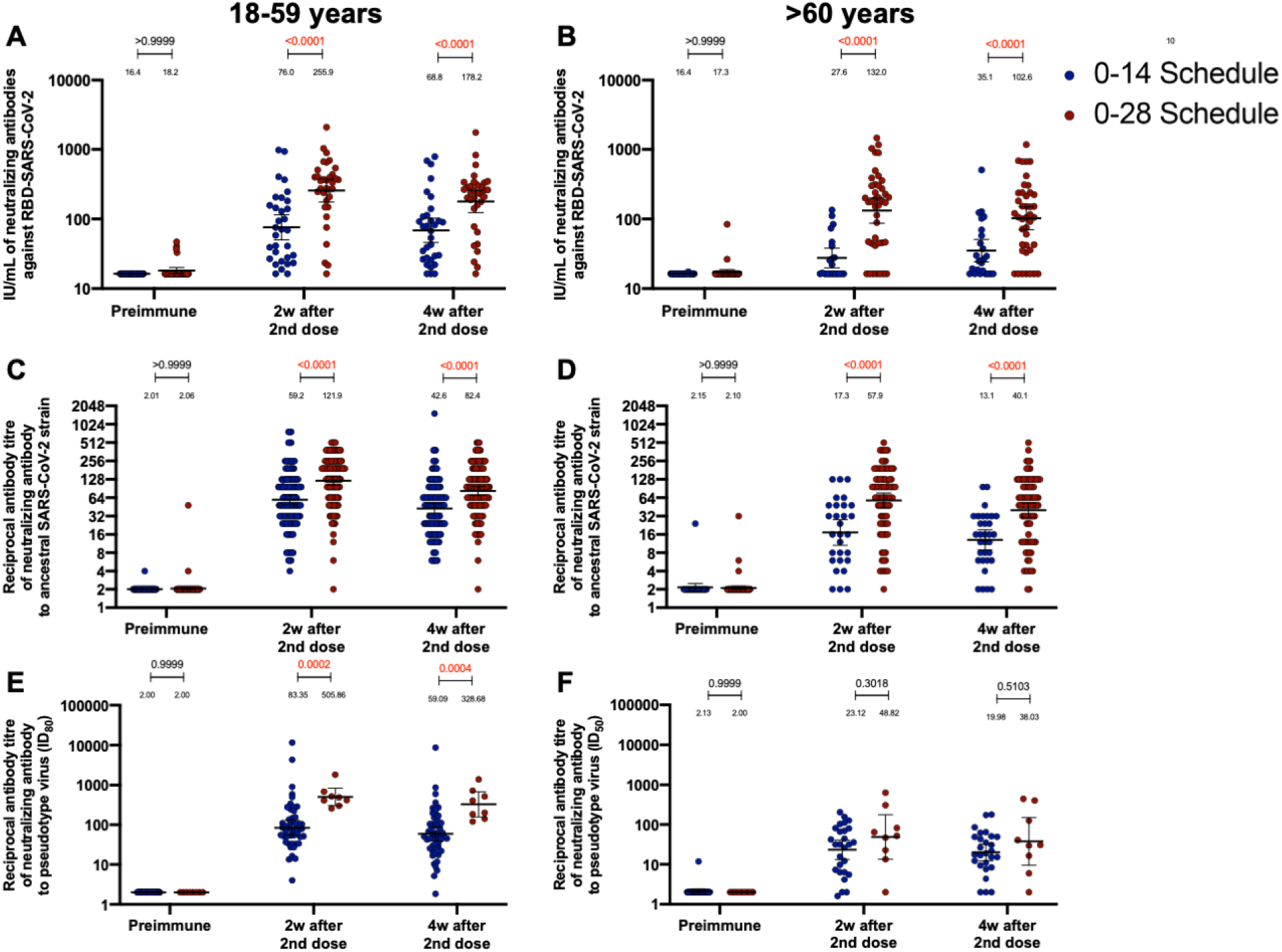
Circulating neutralizing antibodies against SARS-CoV-2 measured through sVNT, cVNT, and pVNT (ID80) in volunteers immunized with CoronaVac aged 18-59 and ≥60 years. Neutralization assays were performed with a surrogate virus neutralization assay (sVNT), which quantifies the interaction between S1-RBD and hACE2 pre-coated on ELISA plates (**A,B**); with a conventional plaque reduction neutralization test (cVNT), which quantifies the cytopathic effect induce in Vero cells as plaques formation for the Ancestral and D614G strains (**C-F**); and with a pseudovirus-based neutralization test (pVNT) (**G,H**). Results were obtained from volunteers aged 18-59 years old (**left panel, A,C,G,E**) and ≥60 years old (**right panel, B,D,F,H**) before immunization (0 days), two weeks after second dose (42 days), and 4 weeks after the second dose (56 days). Data is represented as the reciprocal antibody titer v/s time after the second dose. Numbers above the bars show the international units (IU) or Geometric Mean Titer (GMT), and the error bars indicate the 95% CI. A two-tailed, one-way ANOVA for repeated measures was performed over the Log_2_ of antibody titers, followed by Bonferroni’s multiple comparisons test to compare between schedules. Values above the lines indicate P values. Significant P values are shown in red.

**Figure S4.**
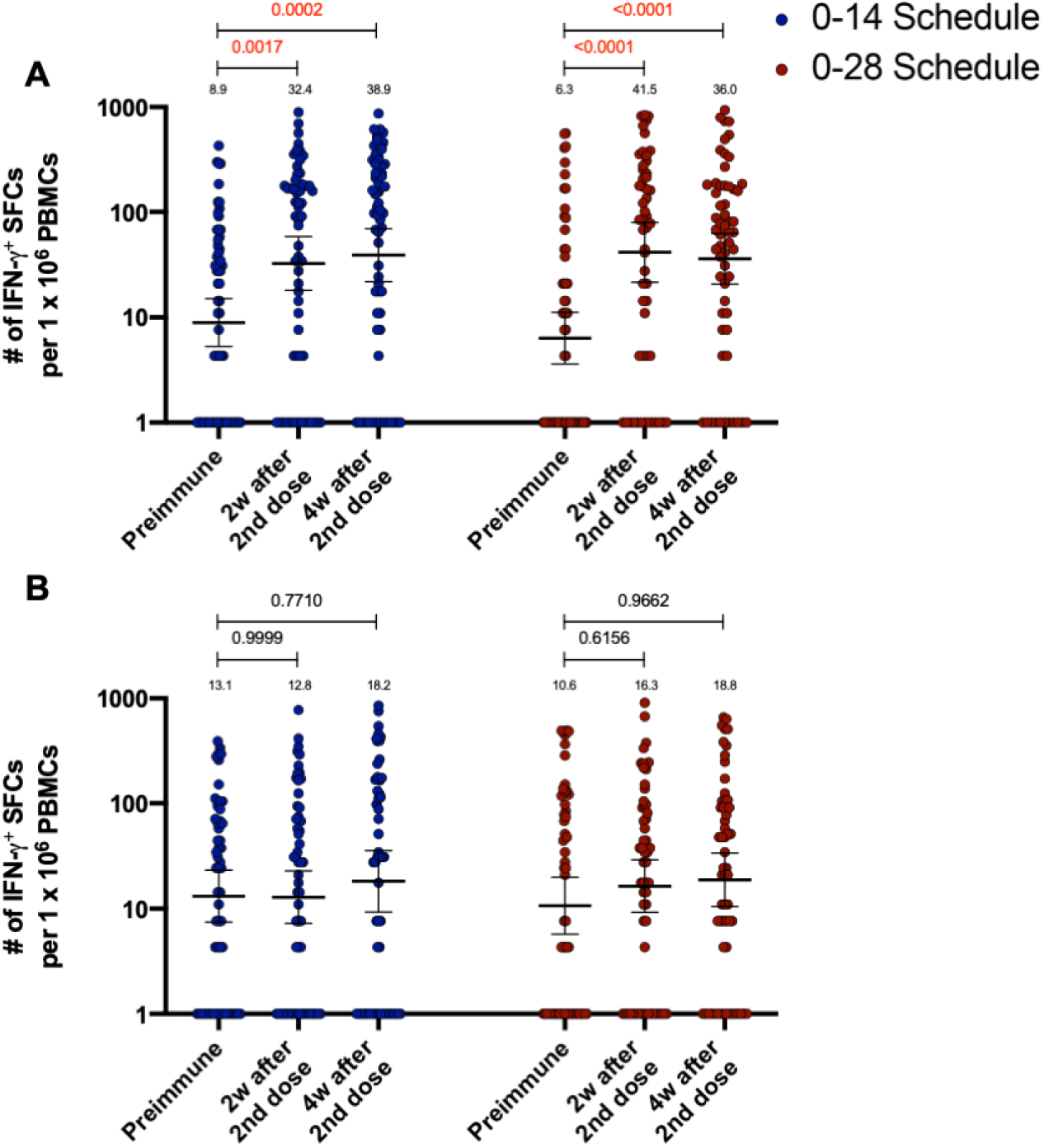
Total number of IFN-γ^+^ SFCs induced upon stimulation with MPs of peptides derived from SARS-CoV-2 proteome. Changes in the secretion of IFN-γ were measured, determined as the number of Spot Forming Cells (SFC) per 1×10^6^ PBMC. Data was obtained upon stimulation of PBMC with MP-S+R (**A**), and upon stimulation of PBMC with MP-CD8A+B (**B**), for 48h in samples obtained before immunization, two weeks after the second dose, and four weeks after the second dose. Data are presented as geometric means (numbers above the bars) and error bars represent the 95% CI. A Mixed-effects two-way ANOVA for repeated measures was performed over the Log_10_ of SFCs, followed by Bonferroni’s multiple comparisons test to compare between schedules. Values above the lines indicate P values. Significant P values are shown in red.

**Figure S5.**
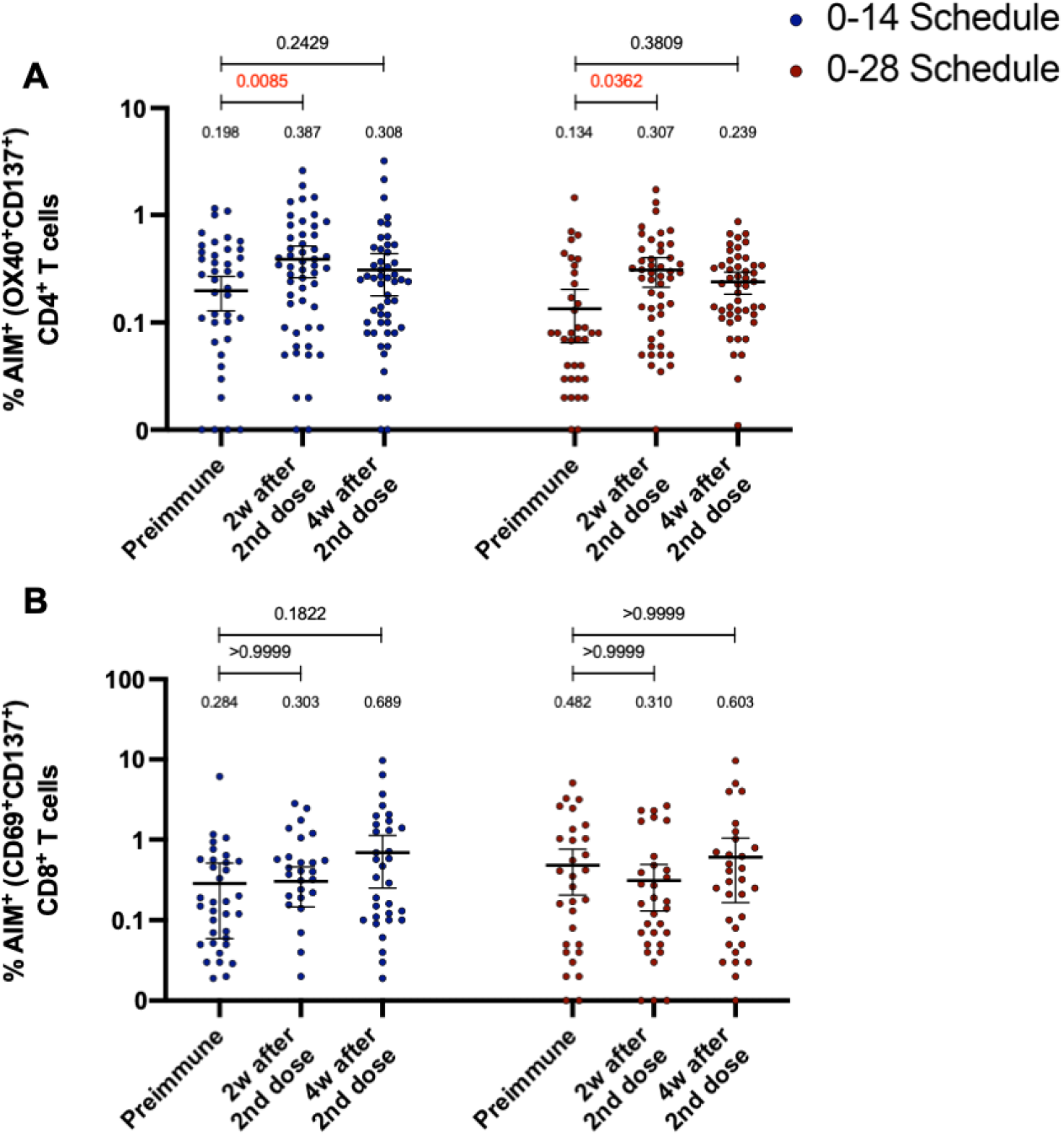
Percentage of AIM^+^ T cells induced upon stimulation with MPs of peptides derived from SARS-CoV-2 proteome in volunteers immunized with CoronaVac. The percentage of activated CD4^+^ (AIM^+^ [OX40^+^, CD137^+^]) and CD8^+^ (AIM^+^ [CD69^+^, CD137^+^]) T cells was determined by flow cytometry, upon stimulation for 24h with MP-S+R (**A**), or MP-CD8A+B (**B**) in samples obtained before the first (preimmune) and second dose, and two and four weeks after the second dose. Data are presented as means (numbers above bars) and error bars represent the 95% CI. Mixed-effects two-way ANOVA for repeated measures was performed, followed by Bonferroni’s multiple comparisons test. Values above the lines indicate P values. Significant P values are shown in red.

**Figure S6.**
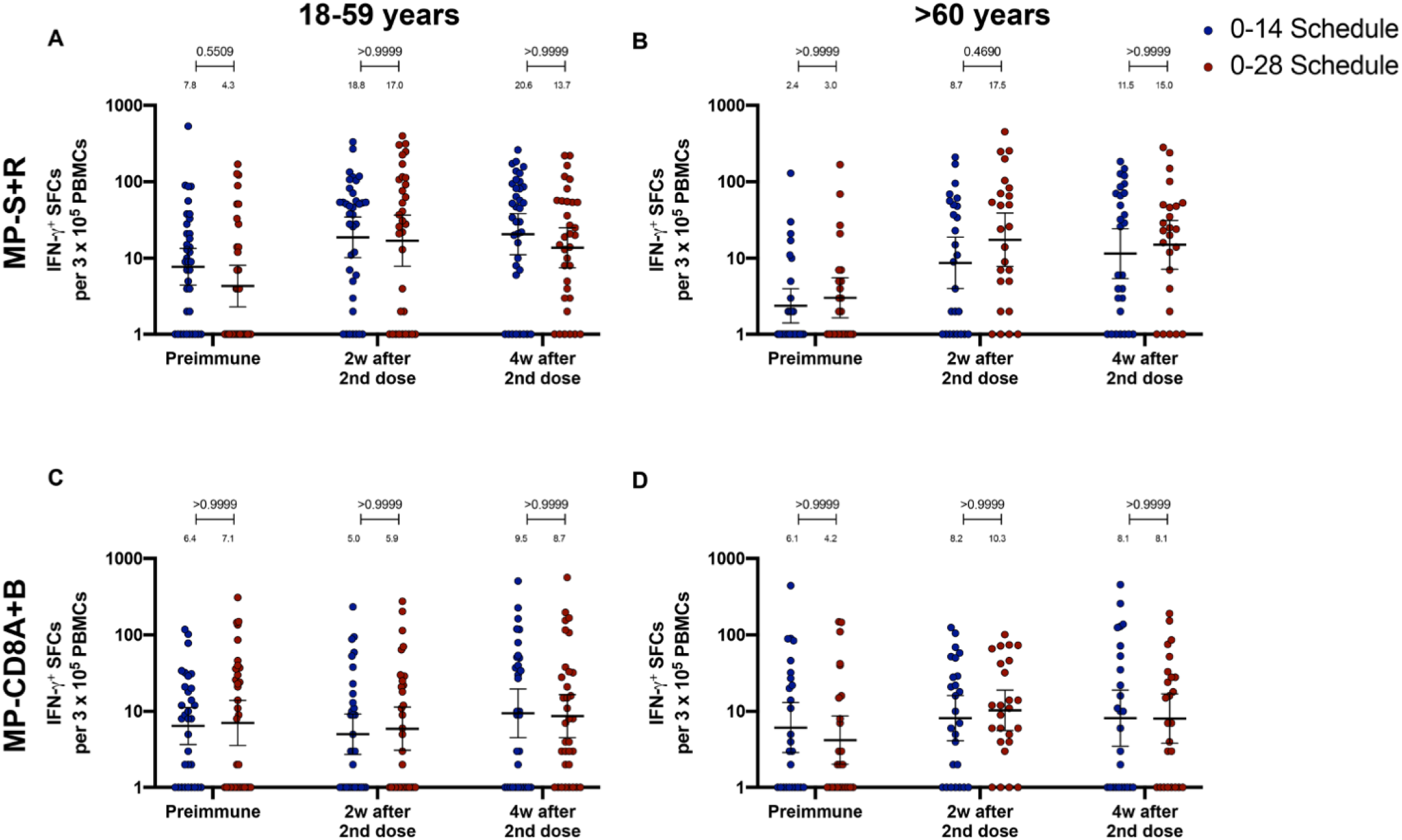
Total number of IFN-γ^+^ SFCs induced upon stimulation with MPs of peptides derived from SARS-CoV-2 proteome in volunteers immunized with CoronaVac aged 18-59 and ≥60 years. Changes in the secretion of IFN**−**γ were measured, determined as the number of Spot Forming Cells (SFC) per 3×10^5^ PBMC. Data was obtained upon stimulation of PBMC with MP-S+R (**A**), and upon stimulation of PBMC with MP-CD8A+B (**B**), for 48h in samples obtained before immunization, two weeks after the second dose, and four weeks after the second dose. All data was normalized for DMSO unspecific stimulation. Data are presented as geometric means and error bars represent the 95% CI. A two-tailed, one-way ANOVA for repeated measures was performed over the Log_10_ of SFCs, followed by Bonferroni’s multiple comparisons test to compare between schedules. Values above the lines indicate P values. Significant P values are shown in red.

**Figure S7.**
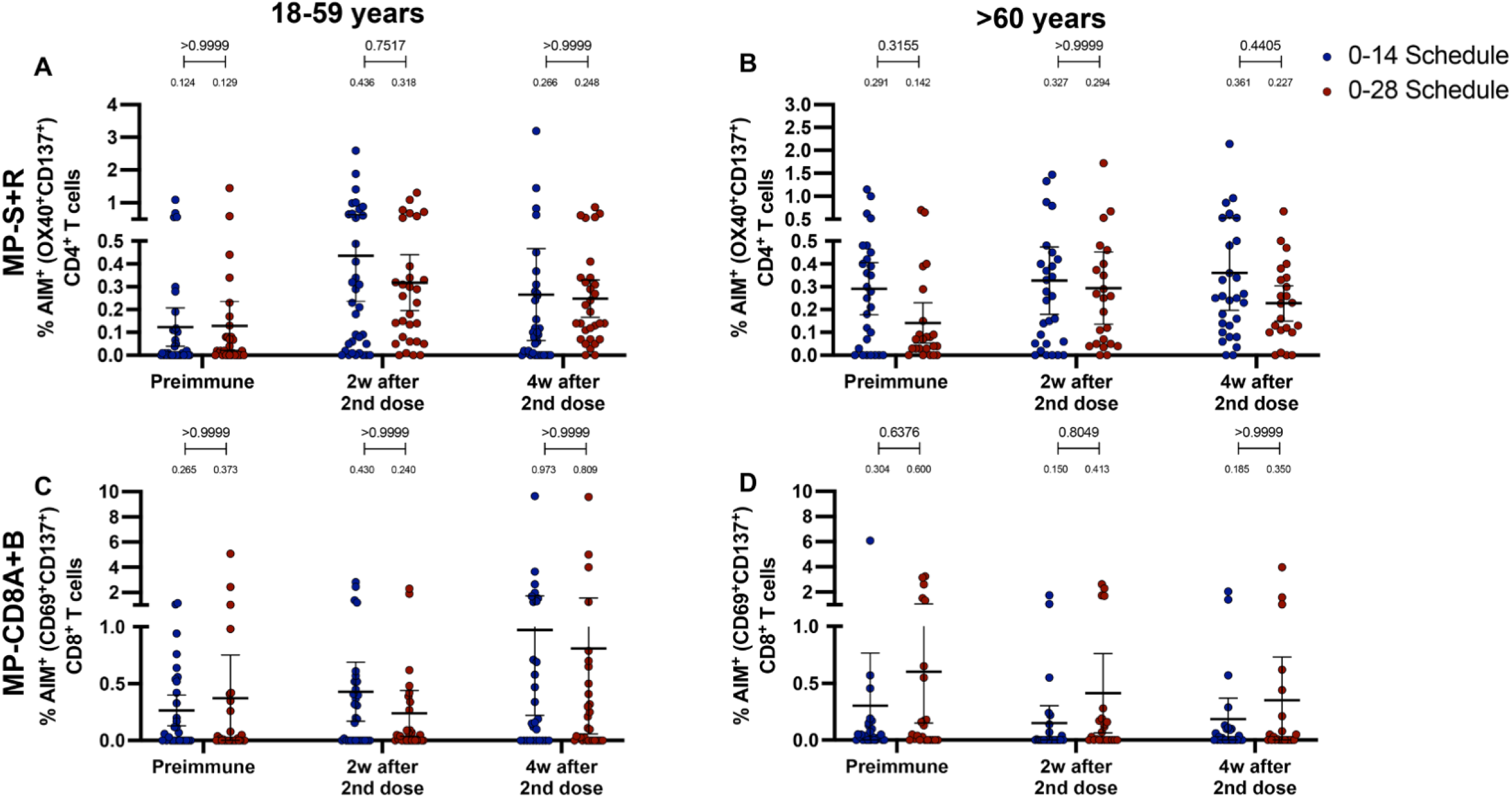
Percentage of AIM^+^ T cells induced upon stimulation with MPs of peptides derived from SARS-CoV-2 proteome in volunteers immunized with CoronaVac aged 18-59 and ≥60 years. The percentage of activated CD4^+^ (AIM^+^ [OX40^+^, CD137^+^]) and CD8^+^ (AIM^+^ [CD69^+^, CD137^+^]) T cells was determined by flow cytometry, upon stimulation for 24h with either MP-S or -R (**A-B**), and with either MP-CD8A or -B (**C-D**) in samples obtained before the first (preimmune) and second dose, and two and four weeks after the second dose. Data are presented as means and error bars represent the 95% CI. A two-tailed, one-way ANOVA for repeated measures was performed, followed by Bonferroni’s multiple comparisons test to compare between schedules. Values above the lines indicate P values. Significant P values are shown in red.

**Figure S8.**
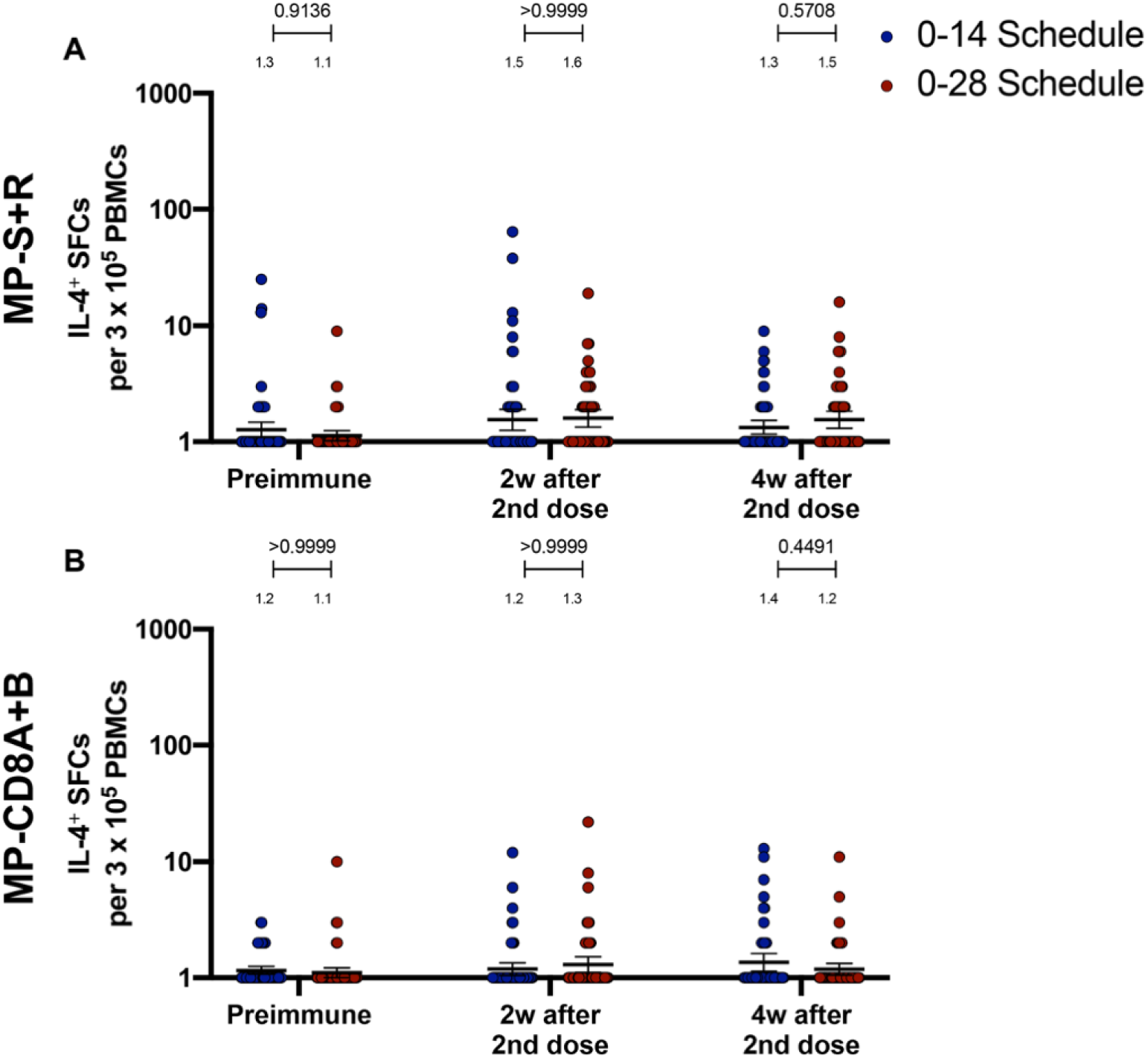
Immunization with CoronaVac in a 0-28 schedule does not induce major IL-4 responses in PBMCs. Changes in the secretion of IL-4 were measured, determined as the number of Spot Forming Cells (SFC) per 3×10^5^ PBMC. Data was obtained upon stimulation of PBMC with MP-S+R (**A**), and upon stimulation of PBMC with MP-CD8A+B (**B**), for 48h in samples obtained before immunization, two weeks after the second dose, and four weeks after the second dose. All data was normalized for DMSO unspecific stimulation. Data are presented as geometric means and error bars represent the 95% CI. A two-tailed, one-way ANOVA for repeated measures was performed over the Log_10_ of SFCs, followed by Bonferroni’s multiple comparisons test to compare between schedules. Values above the lines indicate P values. Significant P values are shown in red.

**Figure S9.**
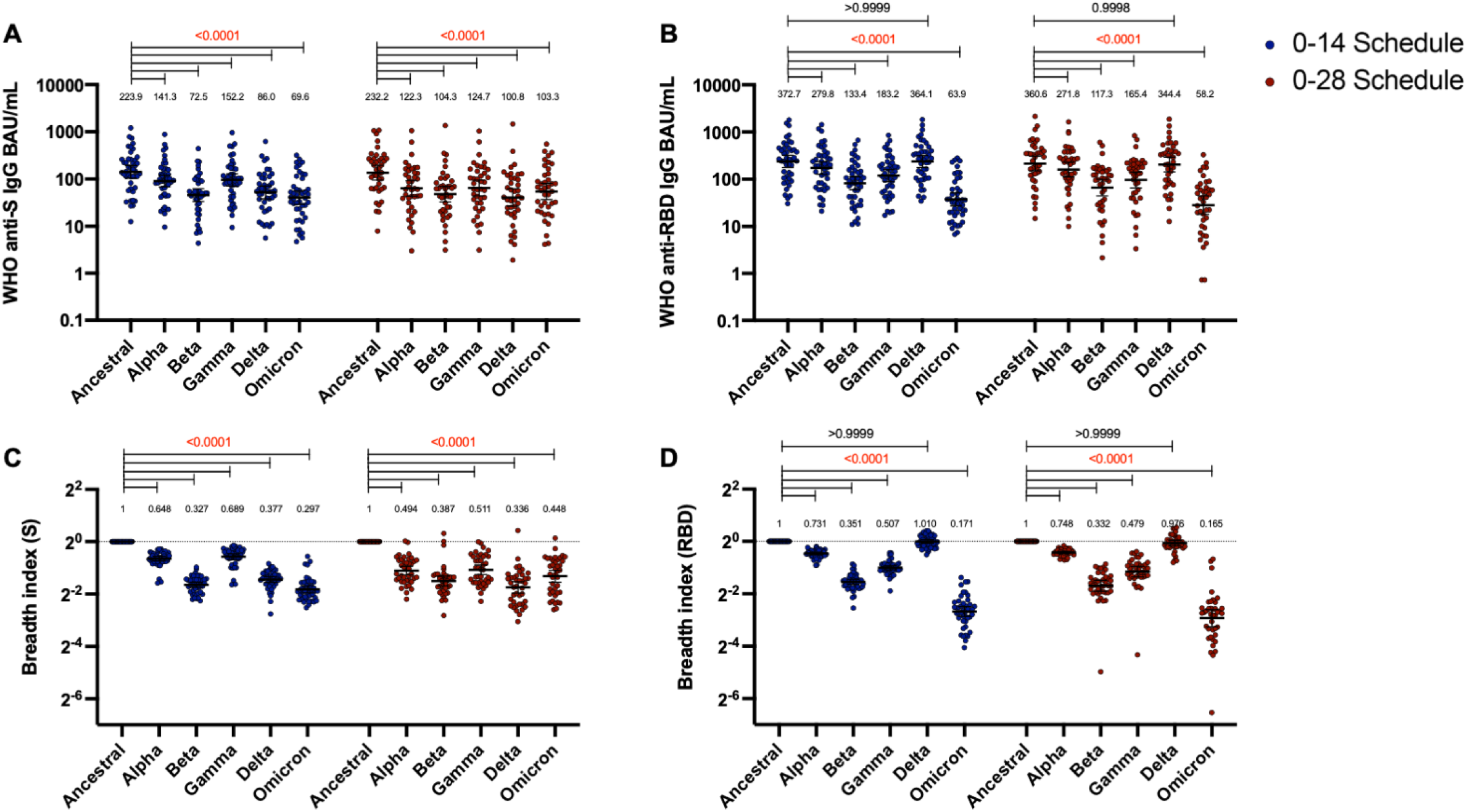
Antibodies against the S and RBD from VOC of SARS-CoV-2 and their breadth indexes are reduced relative to the Ancestral strain, except in the RBD-related parameters for the Delta strain. Antibodies concentrations against the S (**A**) and the RBD (**B**) of different VOC of SARS were evaluated through MSD. Results were obtained from 44 volunteers for the 0-14 schedule and 40 volunteers for the 0-28 schedule, from samples obtained at 4 weeks after the second dose. Data is represented as the reciprocal antibody titer of neutralizing antibody v/s the different VOC evaluated. With these values, a breadth index was calculated for each VOC for anti-S (**C**) and anti-RBD (**D**) antibodies. Numbers above the bars show either the international units (IU) (**A, B**) or the breadh index (**C, D**), and the error bars indicate the 95% CI. Data were analyzed by a Mixed-effect two-way ANOVA, followed by a Bonferroni’s *post hoc* test to compare immunization schedules. Numbers above each bracket represent calculated P values comparing both immunization schedules.

**Figure S10.**
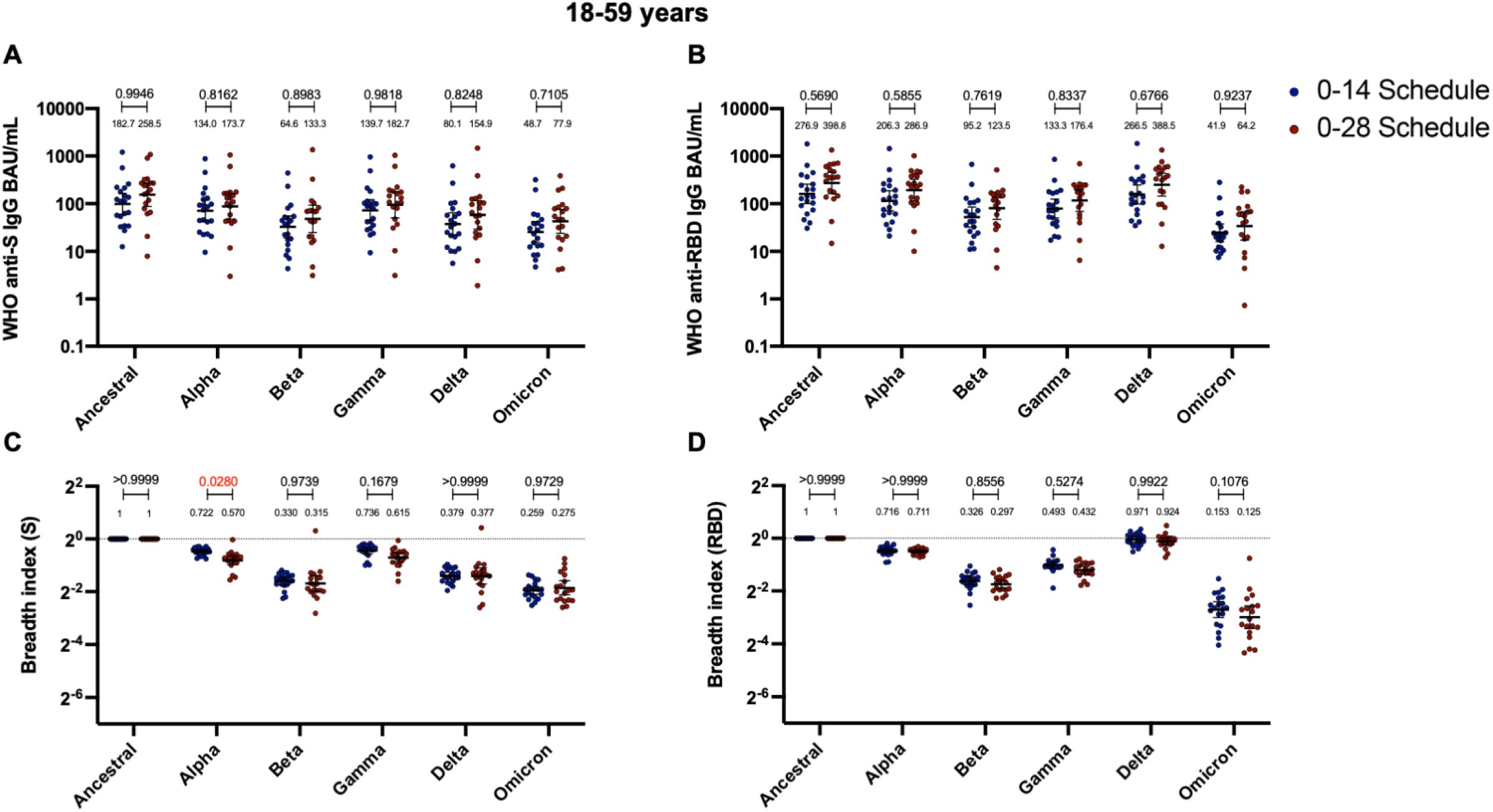
Antibodies against the S and RBD from VOC of SARS-CoV-2 are similar between schedules, while breadth index varies between schedules for the 18-59 years old group. Antibodies concentrations against the S (**A**) and the RBD (**B**) of different VOC of SARS were evaluated through MSD. Results were obtained from 20 volunteers for the 0-14 schedule and 19 volunteers for the 0-28 schedule, from samples obtained at 4 weeks after the second dose. Data is represented as the reciprocal antibody titer of neutralizing antibody v/s the different VOC evaluated. With these values, a breadth index was calculated for each VOC for anti-S (**C**) and anti-RBD (**D**) antibodies. Numbers above the bars show either the international units (IU) (**A, B**) or the breadh index (**C, D**), and the error bars indicate the 95% CI. Data were analyzed by a Mixed-effect two-way ANOVA, followed by a Bonferroni’s *post hoc* test to compare immunization schedules. Numbers above each bracket represent calculated P values comparing both immunization schedules.

**Figure S11.**
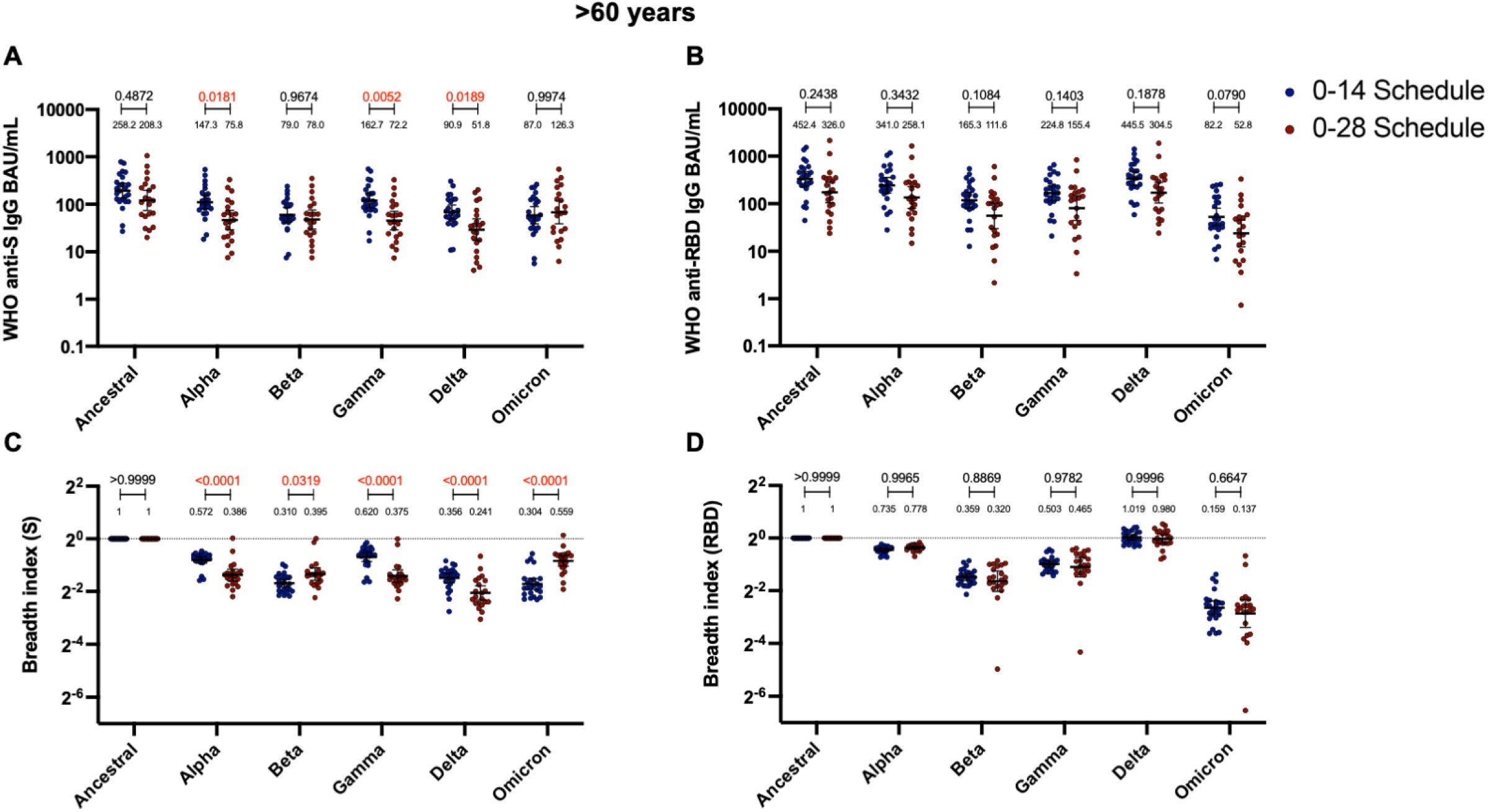
Antibodies against the S from VOC of SARS-CoV-2 and breadth indexes varies between schedules for the >60 years old group, while these parameters for anti-RBD antibodies remain similar. Antibodies concentrations against the S (**A**) and the RBD (**B**) of different VOC of SARS were evaluated through MSD. Results were obtained from 24 volunteers for the 0-14 schedule and 21 volunteers for the 0-28 schedule, from samples obtained at 4 weeks after the second dose. Data is represented as the reciprocal antibody titer of neutralizing antibody v/s the different VOC evaluated. With these values, a breadth index was calculated for each VOC for anti-S (**C**) and anti-RBD (**D**) antibodies. Numbers above the bars show either the international units (IU) (**A, B**) or the breadh index (**C, D**), and the error bars indicate the 95% CI. Data were analyzed by a Mixed-effect two-way ANOVA, followed by a Bonferroni’s *post hoc* test to compare immunization schedules. Numbers above each bracket represent calculated P values comparing both immunization schedules.

